# Rotational perturbation training improves muscle coordination during reactive standing balance in trained and untrained conditions in children with spastic cerebral palsy

**DOI:** 10.64898/2026.09.14.26361313

**Authors:** Jente Willaert, Anja Van Campenhout, Kaat Desloovere, Friedl De Groote

**Affiliations:** Department of Movement Sciences, KU Leuven, Leuven, Belgium; Department of Development and Regeneration, KU Leuven – UZ Leuven, Leuven, Belgium; Department of Rehabilitation Sciences, KU Leuven – UZ Leuven, Leuven, Belgium

**Author notes:** **Corresponding author** Jente Willaert, Gebouw De Nayer (bus 1501, lokaal 02.49), Tervuursevest 101, 3001 Leuven Belgium.

## Abstract

Perturbation training seems a promising tool to improve reactive balance in children with cerebral palsy. However, experimental evidence remains limited. In particular, little is known about how perturbation training alters muscle coordination, limiting insight into the mechanisms underlying training-induced improvements in balance control. Furthermore, it is unclear whether adaptations in balance control generalize to untrained balance tasks. Here, we investigated the effect of rotational perturbation training on reactive balance performance and reactive muscle activity during trained and untrained perturbation conditions in children with cerebral palsy and typically developing children.

We performed a longitudinal intervention study in fourteen children with cerebral palsy and ten typically developing children. Reactive balance was assessed before and after the intervention using toe-up rotational and backward translational perturbations of standing balance. The training intervention consisted of nine sessions over three weeks, with each session including 96 rotational perturbations. Perturbation intensity was adapted to the participant’s performance level. Reactive balance performance was quantified as the ability to stay upright without stepping. Reactive muscle responses were evaluated using the co-contraction index and average muscle activity.

Rotational perturbation training improved the ability of children with cerebral palsy to maintain standing balance without stepping. All children were able to perform more difficult perturbation levels after training. Muscle co-activation decreased after training in response to both trained rotational and untrained translational perturbations in children with cerebral palsy. In addition, average reactive muscle activity in both plantarflexors and dorsiflexors decreased after training in both typically developing children and children with cerebral palsy.

Our findings suggest that reactive balance control can be modified through perturbation training, and that training-induced adaptations generalize to untrained perturbation conditions.

## INTRODUCTION

Balance impairments are common in children with cerebral palsy (CP) [1], [2], [3]. Reactive balance, defined as the ability to maintain balance after an unpredictable disturbance, is needed in daily gross motor skills such as walking. Therefore, it is not surprising that 35% - 55% of children with cerebral palsy falls daily [4]. Perturbation training has emerged as a promising tool to improve reactive balance [5], [6], [7], [8]. However, research on the effect of perturbation training in children with cerebral palsy and the underlying neuromuscular mechanisms is very limited. Previous perturbation training studies using translational perturbations on a moving platform demonstrated improvements in balance recovery and suggested mechanisms such as faster muscle activation, better distal-proximal sequence, and amplitude modulation [7], [8]. Nevertheless, these findings were based on a small sample size (N=6) and mainly focused on a qualitative description of temporal aspects of the postural responses. Therefore, there is a lack of detailed quantitative analysis of changes in muscle coordination and co-activation, hampering the understanding of the underlying mechanisms, the comparison with typically developing peers, and the adaptation to different perturbation intensities. Furthermore, it is unclear whether perturbation training also induces adaptations in balance control that generalize to balance conditions that have not been trained.

Reactive balance, which is often studied by applying support-surface perturbations during standing, is impaired in children with cerebral palsy [2], [3], [9], [10], [11]. When standing balance is perturbed by moving the support surface, children with cerebral palsy step at lower perturbation levels (i.e., smaller or slower platform displacements) [3], [9], [10], [12], [13]. Furthermore, children with cerebral palsy activate their muscles more than typically developing children in response to similar center of mass disturbances [12], [13], they activate their muscles with altered timing and directionality [2], [10], and they have high levels of agonistic-antagonist muscle co-activation [2], [11], [12], [13]. Poor balance control is related to functional disabilities [1].

Perturbation training is a promising tool to improve reactive balance [5], [6], [7], [8]. Perturbations of standing balance can be administered by pushing the subject, changing the compliance of the support-surface, or by moving the support-surface during standing. Support-surface perturbations are particularly suitable as they allow for safe, controlled, and repeatable stimuli. Perturbation training was already successful for improving steady state balance in young healthy adults [14], and to reduce fall risk in older adults with and without fall risk [15], [16], and in neurological populations (e.g., patients with Parkinson’s disease [15] and stroke survivors [17]). However, research on the effect of reactive balance training in children with cerebral palsy is limited.

Perturbation training improves balance performance in children with cerebral palsy but insight in the mechanisms underlying training-induced improvements is limited. In the study of Phillips et al., a balance training protocol based on variable compliance of the support-surface showed promising results in functional balance and fall risk for two children with cerebral palsy [18]. The training consisted of ten sessions in six weeks, and the platform compliance changed with balance performance. However, no analysis of the underlying neuromuscular mechanisms was performed. In the study of Woollacot et al., a balance training protocol based on support-surface backward and forward translational perturbations improved the ability to recover stability in six children with cerebral palsy [7], [8]. The training consisted of five sessions (5 consecutive days) of 100 perturbation trials per session. After training, the center of pressure area reduced, and stability recovered faster with a reduced time to stabilization [7]. Woollacott et al. suggested that the observed gains in stability were driven by improved modulation of muscle responses as evidenced by improved directional specificity (i.e., plantarflexor activity during forward body sway and tibialis anterior activity during backward body sway) and improved amplitude modulation (i.e., reduced muscle co-activation) [8]. Although these studies only had a limited sample size (two and six participants, respectively) and the analyses were mainly qualitative descriptions, these results provide encouraging evidence that reactive balance control remains adaptable in children with cerebral palsy. However, we lack sufficient quantitative analysis of underlying muscle (co-) activation patterns, hampering the understanding of the underlying mechanisms, and we lack a comparison with typically developing peers, making it difficult to determine the difference between pathological motor control and normal developmental variability. Finally, it is unclear whether the benefits of perturbation training are generalizable to untrained balance conditions.

Training of muscle coordination during a specific task might alter muscle coordination during other tasks, but it is unclear if and when such carry-over happens. Animals and humans can learn to modulate their stretch reflex through operant conditioning [19], [20], and such operant conditioning of the stretch reflex improves muscle coordination during walking in spinal cord injured patients [21]. Two studies in older adults found that perturbation training during walking on a treadmill with split-belt accelerations (i.e., inducing slip-wise perturbations) carried over to walking on a slippery surface [22] and faster voluntary stepping responses [23]. While walking on a slippery surface is very similar to treadmill accelerations (i.e., the trained task), voluntary reactive stepping does not challenge reactive balance directly, and therefore provides evidence for carry-over to different tasks. On the other hand, Van Wouwe et al. found that training balance through multi-directional support surface perturbations of standing did not improve reactive balance during walking [24]. While transfer of perturbation training effects to other tasks has been examined in older adults, it remains unknown whether similar carry-over effects also occur in children with cerebral palsy. Carry-over of training effects to other tasks might require adaptations in motor control processes that are common to both tasks [25].

In this study, we evaluated the effect of a rotational perturbation training during stance on (1) balance performance measured as the ability to stay upright without stepping and (2) reactive muscle activity in response to trained rotational perturbations and untrained translational perturbations (carry-over). Toe-up rotational perturbations will stretch the plantarflexors and induce backward body tilt. The plantarflexor stretch reflex will further contribute to backward body tilt. Hence, to successfully control upright posture plantarflexor activity needs to be suppressed, while tibialis anterior activity is needed to restore upright posture. Children with cerebral palsy use more plantarflexor-dorsiflexor co-activation than typically developing children [13]. We hypothesized that rotational perturbation training would reduce co-activation in children with cerebral palsy and thereby improve the ability to withstand perturbations for both the trained rotational perturbations and untrained translational perturbations. Fourteen children with cerebral palsy and ten typically developing children performed a 3-week training intervention. Each of the 9 sessions consisted of 96 rotational perturbations during standing, with a subject-specific difficulty level that was adapted from session to session. Before and after the training intervention, we assessed responses to trained rotational and untrained translational perturbations of standing balance.

## MATERIAL & METHODS

### Subjects

The study was approved by the ethical committee of UZ Leuven/KU Leuven (S65671). Fourteen children with cerebral palsy and ten typically developing children (**Table 1**) participated in this study that consisted of a training intervention (3 weeks) with pre- and post-intervention assessments (**Figure 1**). Children with cerebral palsy were recruited through the cerebral palsy reference center at the university hospital Leuven (Belgium). All patients were diagnosed as having spastic CP by a neuro-pediatrician and met the following inclusion criteria: (1) 5 to 17 years old; (2) Gross Motor Function Classification Scale (GMFCS) I-III; (3) able to stand independently for at least 10 minutes; (4) no orthopedic or neurological surgery in the previous year; and (5) no botulinum neurotoxin injections in the previous six months. Typically developing children were recruited through colleagues and friends and were gender and age matched (group-level) with the children with cerebral palsy (**Table 1**).

**Table 1:**
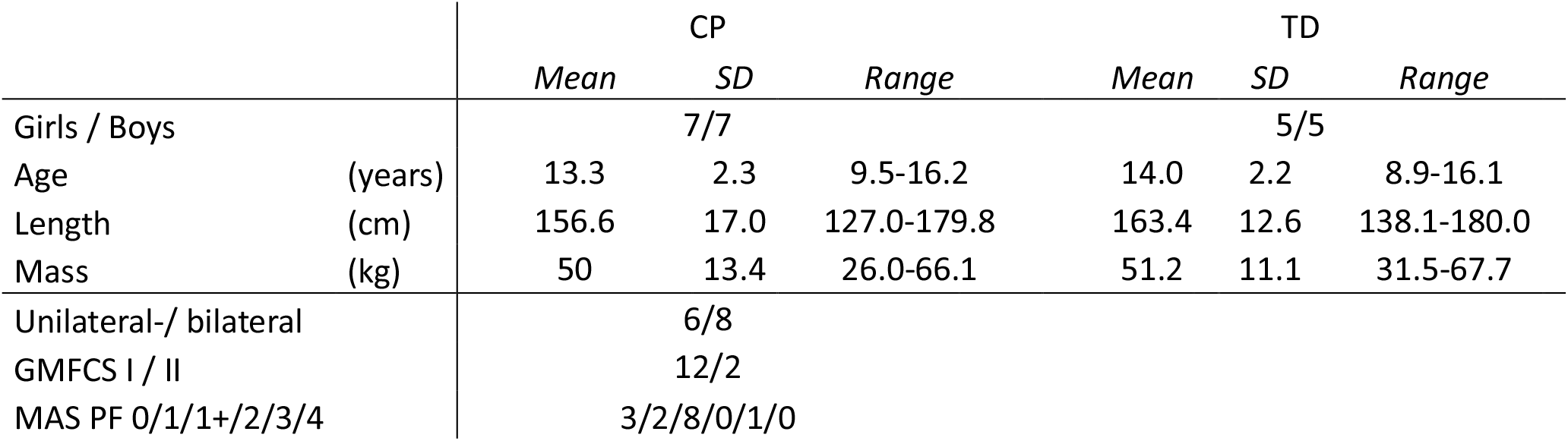
Demographic data of participants (mean, standard deviation, and range)

**Figure 1:**
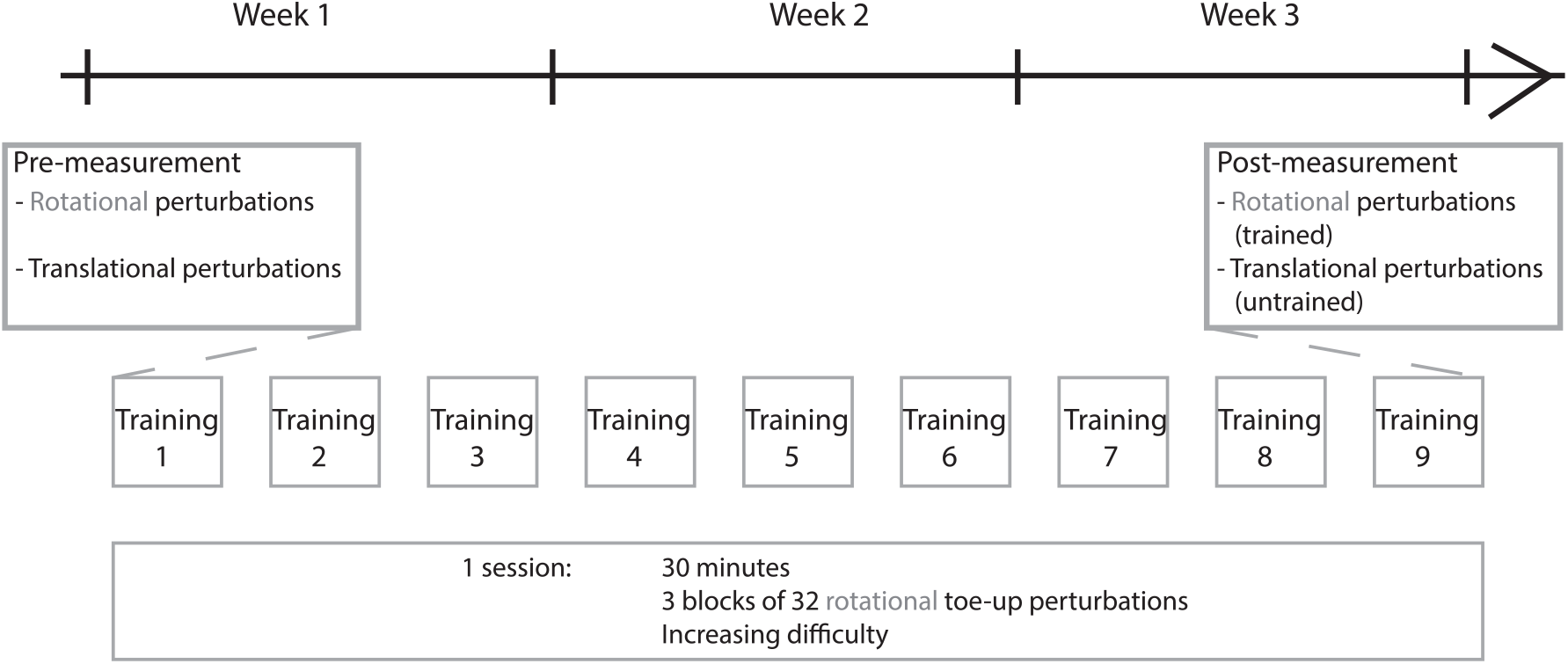
Flowchart of intervention study protocol.

Six children with cerebral palsy were unilaterally involved and eight children were bilaterally involved. Twelve children with cerebral palsy were classified as GMFCS level 1 and two children as GMFCS level 2. Eleven children had spasticity in the gastrocnemius muscle, as indicated by a Modified Ashworth Scale (MAS) higher than zero (**Table 1**). All children were able to stand and walk independently. For the children with cerebral palsy, the most affected leg, based on clinical spasticity scores, was used for further analysis, whereas for typically developing children, the right leg was analyzed.

### Protocol

A Legal representative of the participant and all participants signed an informed consent and informed assent form respectively, before the start of the measurements, according to the principles of the declaration of Helsinki. All measurements and training sessions were performed in the Movement and Posture Analysis Laboratory Leuven (MALL, Belgium).

#### Pre- and post-measurement assessments

The pre- and post-intervention assessments evaluated balance performance and muscle coordination during perturbations of standing balance (**Figure 1**).

Reactive balance was assessed by support-surface perturbations of standing. We applied two different perturbation conditions: (1) combined translational and toe-up rotational perturbations (hereafter called toe-up rotational perturbations), and (2) pure backward translational perturbations (hereafter called backward translational perturbations) on a moveable platform (Caren platform, Motek, The Netherlands) (**Figure 2**). The toe-up rotational perturbations consist of an initial small translational component (max 5° during rotational perturbations compared to min 12.5° in pure translational perturbations, due to the position of the rotation axis of the platform) combined with a big rotational component. These rotational perturbations induced an initial plantarflexor response, followed by a switch to tibialis anterior activity [13]. The backward translational perturbations induced forward body sway with plantarflexor activity needed to maintain balance [12]. Participants were instructed to stand upright and maintain balance without taking a step, unless necessary to avoid falling. When participants needed to take a step, we asked them to return their feet to the starting position, which was marked with tape on the platform. Arm movement was unconstrained. Participants were secured using a safety harness, adjusted to an overhead rail to prevent falling. The protocol consisted of six increasingly difficult perturbation levels (increasing platform displacement, velocity, and/or acceleration) for both perturbation conditions (**Figure 2**). Within each perturbation level, eight perturbations were administered. In the pre- and post-intervention assessments, the participant could proceed to the next level, if balance was maintained without stepping in at least five out of eight trials. If needed, rest was given between the levels.

**Figure 2:**
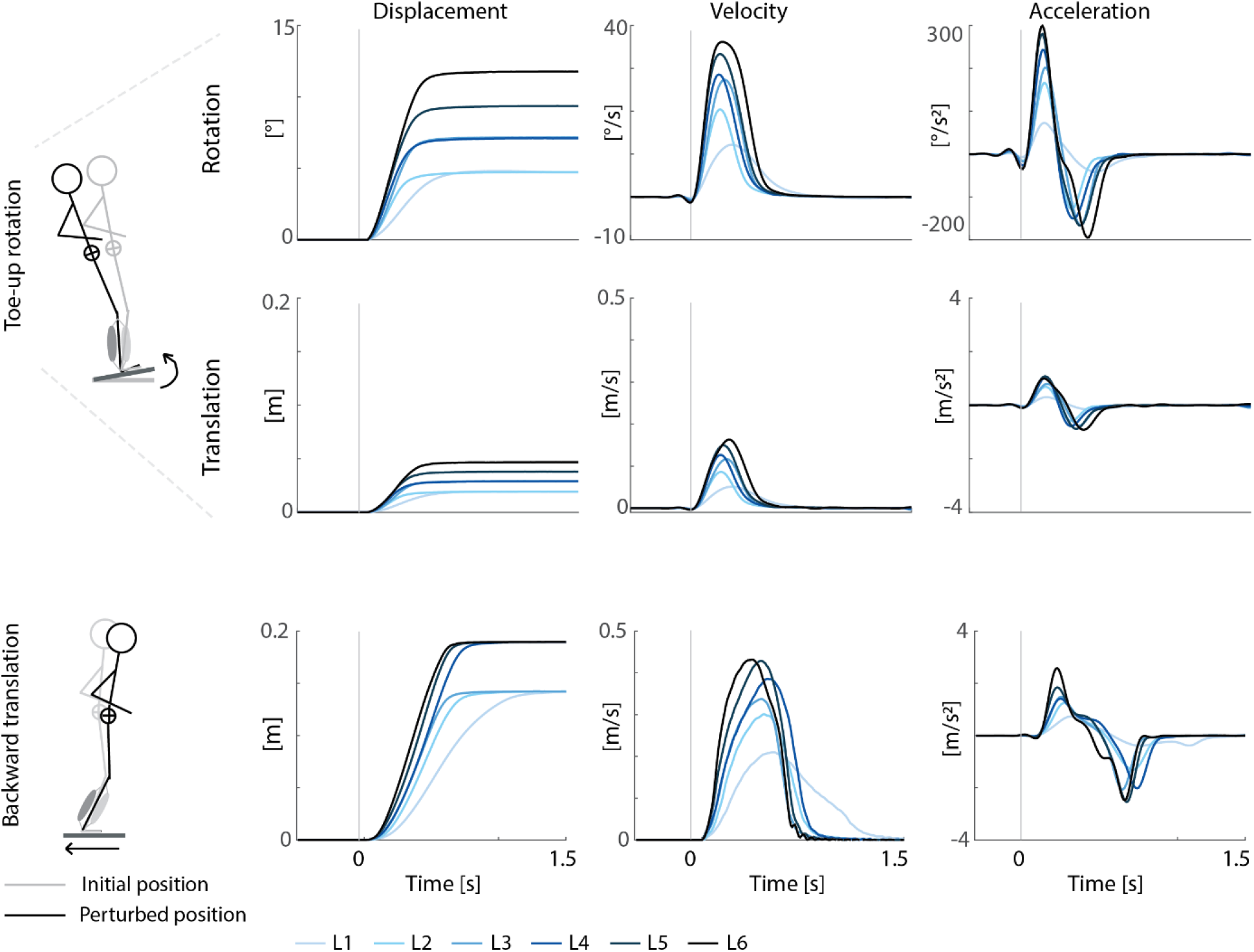
Platform kinematics for the different perturbation levels (L1-L6 in blue) for toe-up rotations (top row) and backward translations (bottom row).

The measurements described here were part of a larger measurement session, more generally assessing joint hyper-resistance and balance during standing and walking.

#### Training protocol

The training intervention (**Figure 1**) consisted of nine sessions (spread over 3 weeks), which took about 30 minutes each. The first and last session immediately followed and preceded respectively the pre- and post-intervention assessments. During one session, we administered three identical blocks of 32 toe-up rotational perturbations. Each block consisted of perturbations with two different difficulty levels (16 trials each). The administered perturbation levels were selected based on the participant’s performance in the previous training session (for training sessions two to nine) and pre-intervention assessment (for the first training session). If the participant was able to maintain balance without stepping in at least 80% of the trials, we continued with a more difficult level in the next training. When the participant was able to perform all levels, we continued the training by combining both lower and higher levels, in order to train the complete range of perturbation levels. An overview of the individual training schemes can be found in **Supplementary material S1, Table S1**. Rest was provided between blocks. Children were asked to maintain balance without taking a step, unless necessary to avoid falling. Only rotational perturbations were trained.

### Data processing and analysis

Trajectories of reflective skin markers (see **Supplementary material S2, Table S2**, for detailed marker placement) and platform movement (3 markers) were captured by 7 infrared Vicon cameras (Vicon, Oxford Metrics, United Kingdom, 100 Hz) during pre- and post-training assessments. Activity of the lateral gastrocnemius (LG), medial gastrocnemius (MG), soleus (SOL), and tibialis anterior (TA) was measured using surface electromyography (sEMG, ZeroWire EMG Aurion, Cometa, Italy, 1000 Hz for CP1-CP11 and TD1-TD7; Trigno Wireless Biofeedback System, Delsys, Natick, USA, 1000 Hz for CP12-CP14 and TD8-TD10). EMG electrodes (Ambu Blue Sensor, Ballerup Denmark for the Cometa system and Avanti Sensor, Delsys for the Delsys system) were placed according to SENIAM guidelines [26].

EMG data was band-pass filtered using a fourth order Butterworth filter between 10 and 450 Hz, followed by signal rectification and low-pass filtering using a fourth order Butterworth filter with 40 Hz cut off. The filtered EMG signal was scaled to the average EMG measured during walking for each measurement session and each participant. We do not expect substantial changes in average muscle activation during walking after balance training. Therefore, average EMG during walking provides a consistent within-participant reference across measurement sessions. EMG parameters for balance were calculated across all non-stepping trials within one perturbation level for each participant.

### Outcome parameters

#### Balance performance

Balance performance was evaluated as the number of levels the participant was able to complete before and after training. A level was considered successful if the participant was able to maintain balance without stepping in at least five out of eight trials.

#### Co-contraction index

We computed the co-contraction index (CCI), a measure of muscle co-activation, as the common area below tibialis anterior and respectively lateral gastrocnemius, medial gastrocnemius, and soleus filtered and scaled EMG trajectories averaged over time (equation 1, [27]) during rotational and translational perturbations of standing balance (**Figure 3**).

**Figure 3:**
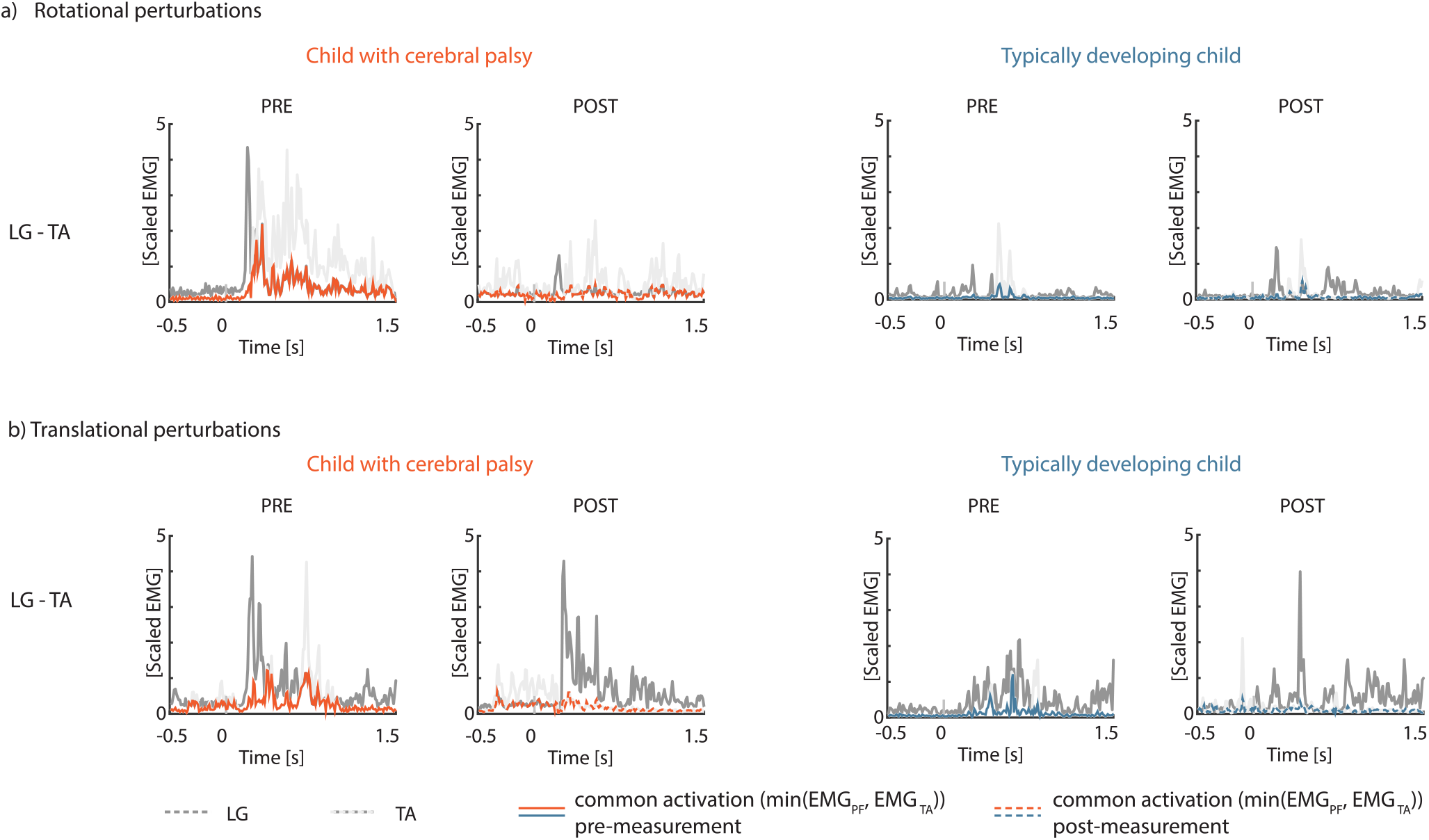
Exemplar EMG trajectories used to compute the co-contraction index (CCI) between the lateral gastrocnemius (LG) and tibialis anterior (TA) for perturbation level 3 (a) rotational perturbation and (b) translational perturbation before (full line) and after (dotted line) training for a child with cerebral palsy (left, in orange) and a typically developing child (right, in blue). Dark and light grey traces represent exemplar trajectories of respectively lateral gastrocnemius and tibialis anterior activity for a single trial of level 3. The co-contraction index is calculated as the time average of the minimum of both signals at every point in time.

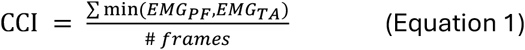

with PF referring to lateral gastrocnemius, medial gastrocnemius, or soleus; and # frames the number of data points in the analyzed time interval. The CCI was calculated for each trial over an interval from 0.5s before perturbation onset until 1.5s after perturbation onset [13]. Trials where the participant took a step were excluded. The average CCI across all non-stepping trials was calculated for each level.

#### Average muscle activity within three time zones

We computed average muscle activity for lateral and medial gastrocnemius, soleus, and tibialis anterior in three time bins (**Figure 4**) during rotational and translational perturbations of standing balance. Reactive muscle activity within each time bin was computed as the average filtered and scaled EMG within the time bin from which baseline activity (average EMG during 100ms preceding perturbation onset) was subtracted. The first time bin (Z1) started at platform onset and ended 150 ms later (**Figure 4, in blue**). During this first time bin, we observed only little muscle activity in both perturbation conditions, possibly due to the slow movement onset of the platform in combination with neural transmission delays. The second time bin (Z2) lasted from 150 ms to 250 ms after platform onset (**Figure 4, in yellow**). During the second time bin, we observed the first peak in plantarflexor activity (LG, MG, and SOL) during rotational perturbations (**Figure 4a**) [13], and backward translational perturbations (**Figure 4b**) in typically developing children [12]. The third time bin (Z3) lasted from 250 ms to 400 ms after perturbation onset (**Figure 4, in red**). During time bin 3, we observed a switch between plantarflexor and tibialis anterior activity in response to rotational perturbations (**Figure 4a**) [13]. For the translational perturbations, we saw a decrease in plantarflexor activity and only limited tibialis anterior activity in typically developing children (**Figure 4b**) [12]. We compared muscle activity within each time bin between pre- and post-intervention assessments.

**Figure 4:**
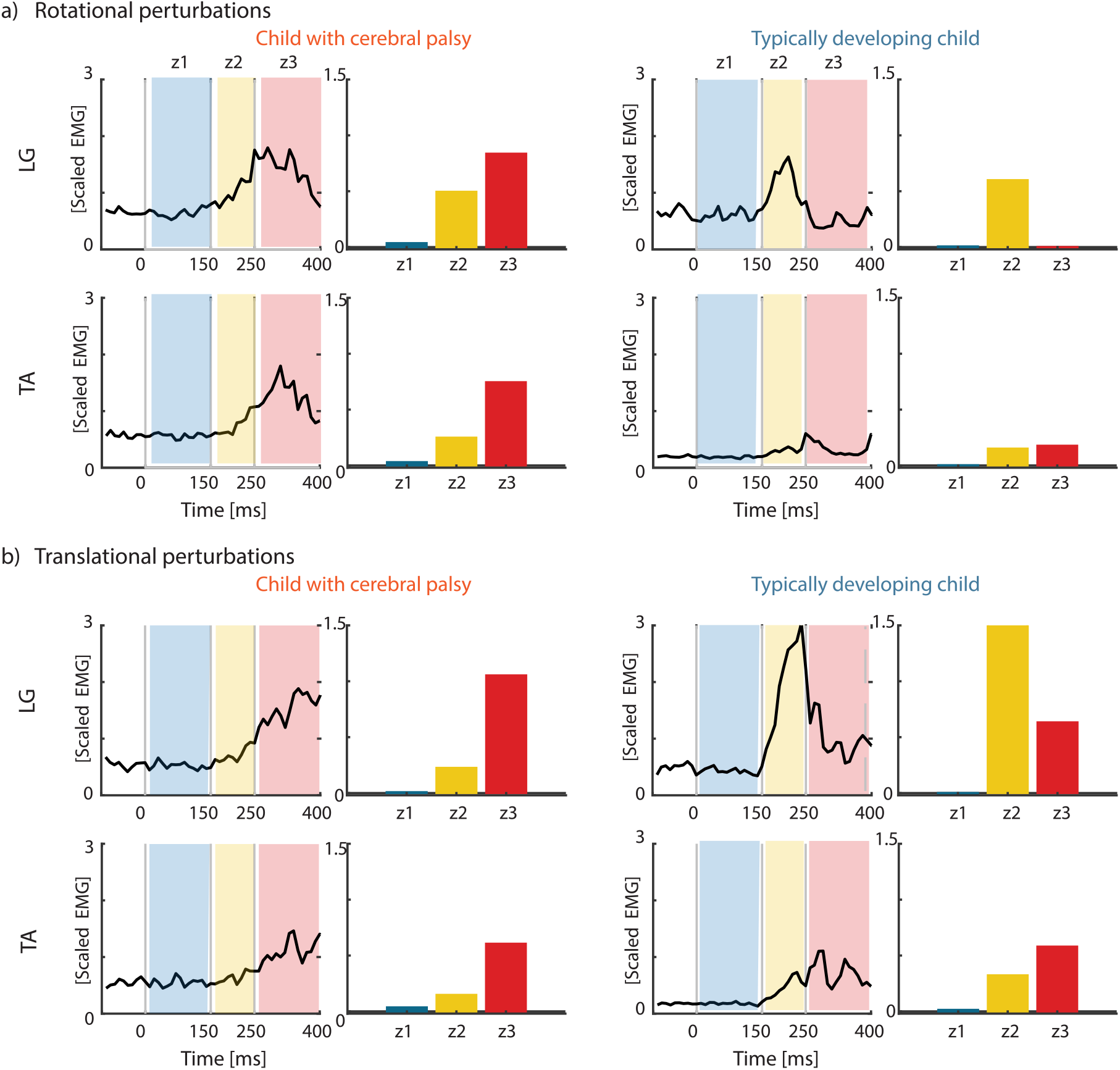
Exemplar EMG trajectories and average muscle activity in each time bin (z1-z3) for perturbation level 2 (a) rotational perturbation and (b) translational perturbation for a child with cerebral palsy (left) and a typically developing child (right). Muscle activations are plotted as a function of time with indication of time bins (colored areas). Bars represent average muscle activity within the indicated time bin.

### Statistics

All statistical analyses were performed using SAS Studio (SAS Institute Inc., Cary, USA).

The co-contraction index was compared between pre- and post-measurement sessions for children with cerebral palsy and typically developing children using a linear mixed model with three fixed effects: (1) time (pre-training vs. post-training), (2) group (CP vs. TD), and (3) level (level 1-6) for each muscle separately. We investigated the effect of time, the effect of group, and the interaction between time and group.

Differences in average muscle activity were compared between pre- and post-measurement sessions for children with CP and TD children using a linear mixed model with three fixed effects: (1) time (pre-training vs. post-training), (2) group (CP vs. TD), and (3) level (level 1-6) for each muscle and time bin separately. We investigated the effect of time, the effect of group, and the interaction between time and group.

## RESULTS

Data were screened for quality prior to analysis. A total of 18 observations were excluded due to technical problems with the EMG system. Excluded observations were distributed across muscles and timepoints (LG: n = 5; MG: n= 6; SOL: n = 2; TA: n = 5).

### Balance performance

Rotational perturbation training improved the ability of children with cerebral palsy to control standing balance without stepping (**Figure 5**).

**Figure 5:**
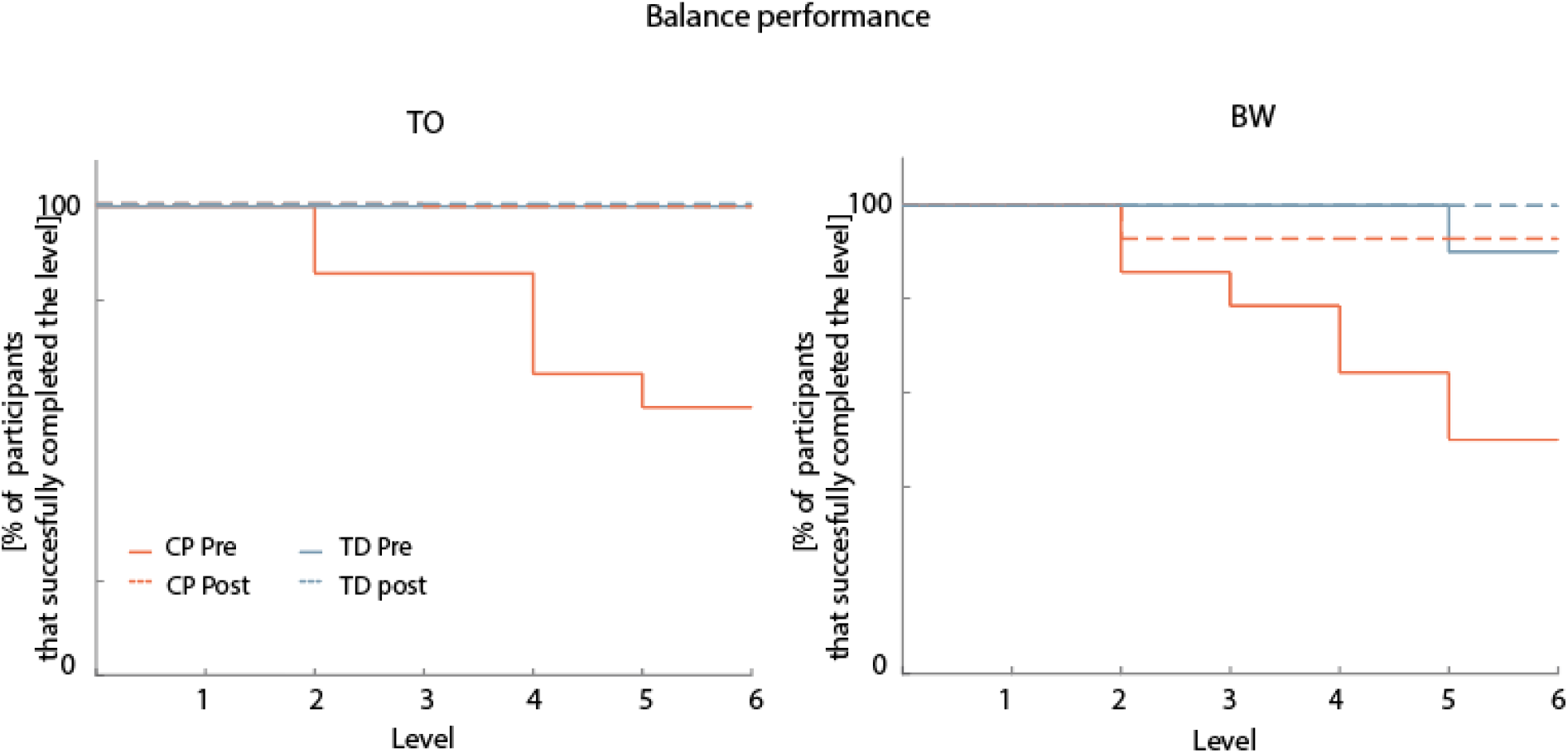
Balance performance before (full line) and after (dotted line) training for toe-up rotational perturbations (TO – left) and backward translation perturbations (BW – right). Percentage of children who were able to withstand perturbations of a certain difficulty level without stepping on more than three out of eight perturbations. Children with cerebral palsy (CP) in orange. Typically developing (TD) children in blue.

All typically developing children were able to perform all rotational perturbation levels before and after training. One typically developing child was not able to perform level 6 of the translational perturbations before training, but all children were able to perform all translational perturbation levels after training.

Only eight children with cerebral palsy were able to complete all rotational perturbation levels before training, one child completed levels 1-5, three children completed levels 1-4, and two children only completed levels 1-2, while all children were able to complete all rotational perturbation levels after training. Seven children with cerebral palsy were able to complete all translational perturbation levels before training, two children completed levels 1-5, two children completed levels 1-4, one child completed levels 1-3, and two children only completed levels 1-2 before training. All children, except one, were able to complete all levels after training. One child was only able to perform levels 1-2 before and after training.

### Co-contraction index

Muscle co-activation decreased after perturbation training for both the trained rotational and untrained translational perturbations in children with cerebral palsy. In typically developing children, co-activation generally remained unchanged after training, except for a decrease in SOL–TA co-activation during the rotational perturbations after training (**Figure 6, Supplementary Material S3. Table S3-S4**). Children with cerebral palsy had higher co-activation before and after training than typically developing children (**Supplementary Material S3. Figure S1-S2, Table S3-S4**).

**Figure 6:**
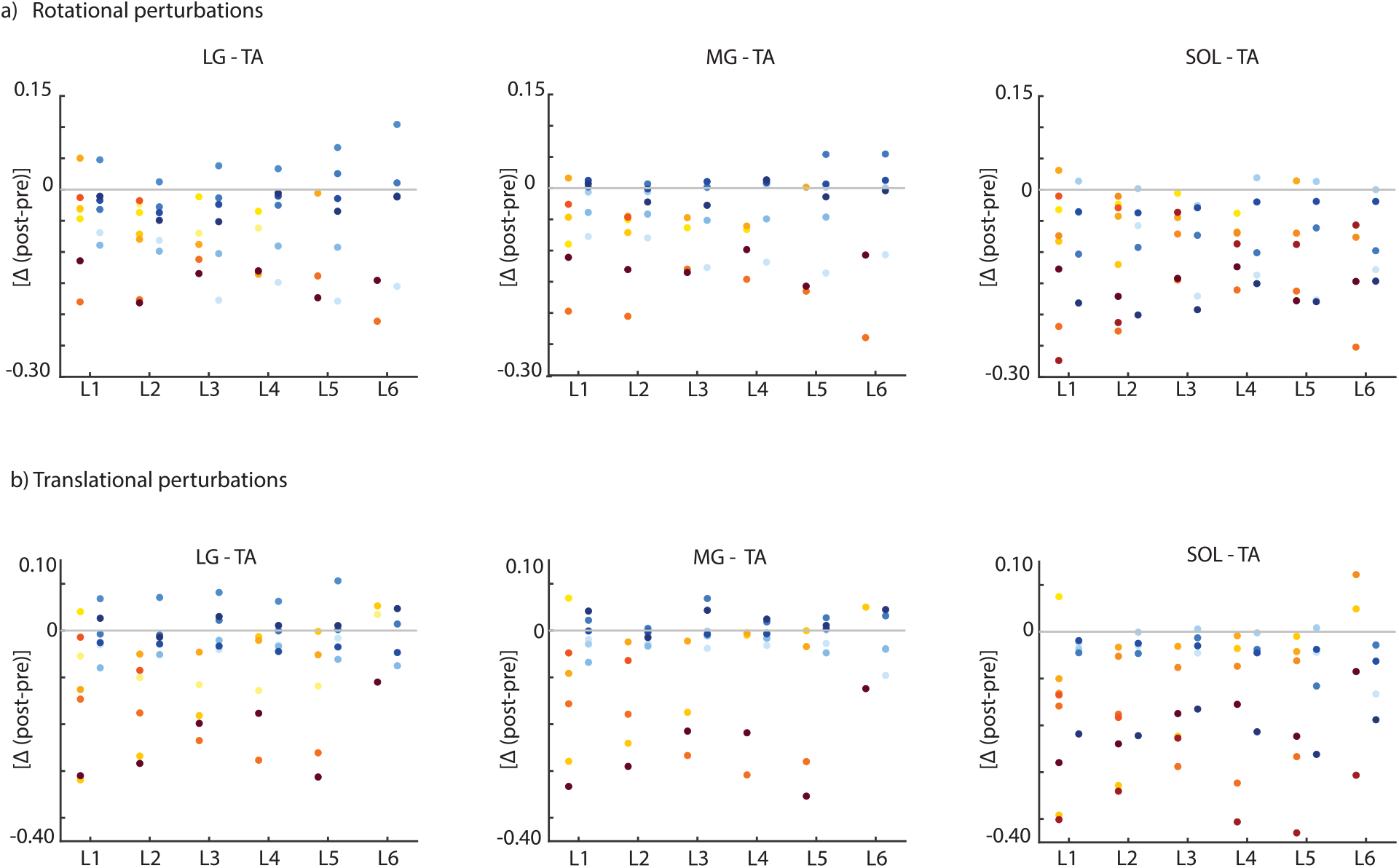
Change (post-measurement – pre-measurement) in co-contraction index between plantarflexors (lateral gastrocnemius (LG), medial gastrocnemius (MG), soleus (SOL)) and tibialis anterior (TA) for (a) toe-up rotational perturbations and (b) backward translational perturbations. Children with cerebral palsy (CP) in red/yellow colors and typically developing (TD) children in blue. Negative values indicate a decrease in co-activation after training.

**Figure 7:**
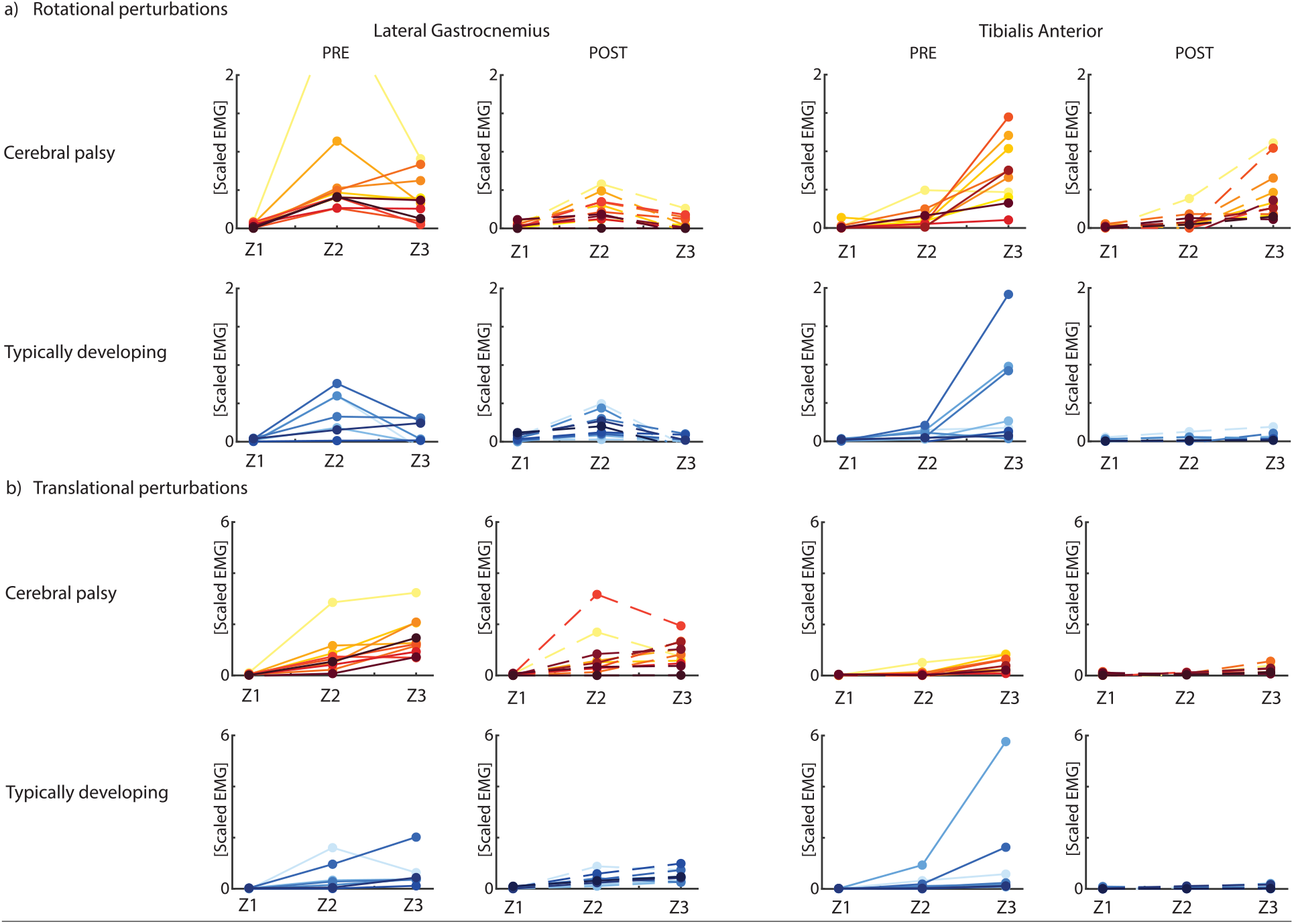
Average normalized EMG for three time bins (Z1-Z3) for level 2 of the rotational perturbations (a – top panel) and translational perturbations (b - bottom panel) pre-(full line) and post (dotted line) intervention. Children with cerebral palsy (CP) in red/yellow colors (top row of each panel) and typically developing (TD) children in blue (bottom row of each panel). Results for all muscles and all levels can be found in **Supplementary Material S4, figures S3-11, Table S5**.

#### Rotational perturbations

The co-contraction index for LG-TA (p=0.046, average difference (post-pre) across all levels (Δ) = -0.076) and SOL-TA (p=0.027, Δ=-0.092) decreased after the training intervention in children with cerebral palsy. No changes were observed for the MG-TA co-contraction index (p=0.224, Δ=-0.068).

Only the co-contraction index for SOL-TA (p=0.005, Δ=-0.068) decreased after the training intervention in typically developing children. No changes were observed for LG-TA (p= 0.119, Δ=-0.044) and MG-TA (p=0.934, Δ=-0.016).

Children with cerebral palsy had higher co-contraction in response to rotational perturbations pre and post training compared to typically developing children for all muscle pairs (LG-TA: p= 0.006; MG-TA: p<0.0001; SOL-TA: p<0.001).

#### Translational perturbations

The co-contraction index for all muscle pairs decreased after the training intervention in children with cerebral palsy (LG-TA: p=0.027, Δ=-0.089; MG-TA: p=0.008, Δ=-0.087; SOL-TA: p=0.018, Δ=-0.127).

No change in co-contraction index for all muscle pairs was observed after the training intervention in typically developing children (LG-TA: p=0.97, Δ=-0.023; MG-TA: p=0.99, Δ=-0.006; SOL-TA: p=0.94, Δ=-0.038).

Children with cerebral palsy had higher co-contraction in response to translational perturbations pre and post training compared to typically developing children for all muscle pairs (LG-TA: p= 0.001; MG-TA: p=0.001; SOL-TA: p<0.001).

### Average muscle activity

After training, both typically developing children and children with cerebral palsy controlled posture with less muscle activity in both plantarflexors and dorsiflexors.

#### Rotational perturbations

Before the training intervention, plantarflexor activity of typically developing children increased from time bin 1 to time bin 2, followed by a decrease from time bin 2 to time bin 3. TA activity increased slightly from time bin 1 to 2, followed by a larger increase from time bin 2 to time bin 3.

After the training intervention, the amplitude of muscle activity for LG (p = 0.011), MG (p < 0.0001), and TA (p = 0.009) decreased in time bin 2, and MG (p = 0.031) and TA (p < 0.0001) activity decreased in time bin 3.

Before the training intervention, plantarflexor activity of children with cerebral palsy increased from time bin 1 to time bin 2, and this increase was larger than in typically developing children. In contrast to typically developing children, plantarflexor activity did not always, or to a lesser extent, decrease from time bin 2 to time bin 3 in children with cerebral palsy. TA activity increased slightly from time bin 1 to 2, and this increase was larger than in typically developing children. Also, the increase in TA activity from time bin 2 to 3 was larger in children with cerebral palsy than in typically developing children.

After the training intervention, the amplitude of muscle activity for LG (p = 0.003) and SOL (p = 0.002) decreased in time bin 2, and LG (p < 0.0001), MG (p = 0.019), and SOL (p = 0.05) activity decreased in time bin 3. Furthermore, the variability across children with cerebral palsy decreased after training.

#### Translational perturbations

Before the training intervention, plantarflexor activity of typically developing children increased from time bin 1 to time bin 2, followed by a similar level of activity in time bin 3 than in time bin 2. TA activity did almost not change from time bin 1 to time bin 2, followed by an increase in activity from time bin 2 to time bin 3.

After the training intervention, the amplitude of muscle activity in response to translational perturbations did not change for typically developing children.

Before the training intervention, plantarflexor activity of children with cerebral palsy increased from time bin 1 to time bin 2, followed by a further increase from time bin 2 to time bin 3. Both the increase from time bin 1 to time bin 2 and from time bin 2 to time bin 3 were larger than in typically developing children. TA activity increased a little from time bin 1 to time bin 2, followed by a larger increase from time bin 2 to time bin 3, suggesting muscle co-activation between plantarflexors and TA. The increase from time bin 2 to time bin 3 was larger in children with cerebral palsy than in typically developing children.

After the training intervention, the amplitude of muscle activity in response to translational perturbations for MG (p = 0.001), SOL (p = 0.010), and TA (p = 0.0001) activity decreased in time bin 3 for children with cerebral palsy.

## DISCUSSION

Perturbation training did improve reactive balance performance (i.e., more difficult perturbations without stepping and did alter the muscle coordination strategy leading to less muscle co-activation in response to perturbations of standing in children with cerebral palsy. Our results suggest that muscle coordination underlying reactive balance control is modifiable in children with cerebral palsy. Although impaired muscle coordination and excessive co-activation are well-known impairments of postural control in cerebral palsy [2], [3], [11], rehabilitation interventions rarely target these neuromuscular deficits directly. The observed reduction in co-activation here supports the potential of perturbation-based interventions to modify underlying motor control strategies. Muscle coordination was improved in both trained rotational and untrained translational perturbations, suggesting that the learned modifications (i.e., decreased muscle co-activation) can be transferred to other perturbation conditions. This suggests that the children did not merely learn a condition-specific response. Instead, they may have gained adaptations in more general reactive balance control mechanisms. Future studies should further investigate the extent to which perturbation training can induce long-lasting changes in neuromuscular control and how this clinically translates into meaningful improvements in balance and daily life activities.

Children with cerebral palsy adapted their muscle coordination to maintain balance through training in a way that reduced the required muscle effort. Increased joint stiffness, due to higher muscle co-activation, may help balance in response to translational perturbations, but not to rotational perturbations. After rotational perturbation training, both plantarflexor and tibialis anterior activity decreased in children with cerebral palsy and typically developing children, yielding increased joint compliance. Suppressing plantarflexor and tibialis anterior activity enables the decoupling of feet and platform movement from the movement of the rest of the body, which might help to keep the body upright while the feet are rotated. In addition, after training, children with cerebral palsy decreased plantarflexor activity when the body sways backward and plantarflexor activity is unfavorable for balance control (time bin 3). This suggests that children with CP were better able to switch between plantarflexor and tibialis anterior activity after training. This is further supported by the decreased CCI for rotational perturbations after training. This improved muscle coordination (i.e., better switch between plantarflexors and tibialis anterior activity and decreased co-activation) might be the reason for improved balance performance, since all children with cerebral palsy were able to perform all rotational perturbations levels without stepping after – but not before - training.

Whereas most perturbation-based training studies used translational perturbations, we used rotational perturbations to target muscle co-activation. During rotational perturbations, muscle co-activation will couple body movement to platform movement resulting in body tilt. Hence, high muscle co-activation, and therefore high joint stiffness, will not be beneficial during rotational perturbations. It might be worthwhile to investigate whether rotational perturbation training induces larger reductions in muscle co-activation than translational perturbation training. Woollacott et al. (2005) already demonstrated that translational perturbation training can improve reactive balance control in children with cerebral palsy and that there are changes in muscle coordination. Our study provides further evidence that perturbation training improves balance performance and hence that reactive balance performance is susceptible to training in cerebral palsy [8]. Patient populations differed between studies. In our study, the cohort was larger (14 versus six participants), providing greater confidence in the observed effects. In addition, our participants had milder motor impairments and nine out of 14 children were older than the participants in Woollacott et al., showing that reactive balance is still susceptible to training in older children. Whereas Woollacott et al. primarily relied on qualitative descriptions of individual muscle activation characteristics, such as onset timing, directional specificity, and distal-to-proximal sequencing, the present study quantitatively evaluated changes in neuromuscular coordination through measures of muscle co-activation. This provides additional insight into the mechanisms underlying improved reactive balance performance. Finally, whereas Woollacott et al. focused exclusively on the trained perturbation paradigm, our study demonstrated improvements in reactive balance for untrained perturbation condition as well.

The reduced muscle co-activation and better performance during untrained translational perturbations suggest that increased co-activation is not a useful compensation to withstand translational perturbations. Children with cerebral palsy often have increased muscle co-activation during translational perturbations of standing [8], [11], [12], [29]. Increased muscle activation leads to increased joint stiffness. This increased stiffness might aid standing balance control in response to translational perturbations by resisting movement of the body with respect to the feet, and can therefore be used as a compensation strategy when balance is impaired. However, the reduced co-activation observed during translational perturbations after training in combination with better performance suggests that a less stiff (i.e., less muscle co-activation) response might be better for balance control. In a previous study, we found that children with cerebral palsy that had a more stiff response to translational perturbations, characterized by a smaller CoM excursion, were not able to perform the more difficult perturbation levels [12], confirming that a stiffer response is not necessarily better for balance control. Furthermore, as mentioned above, a reduction in muscle co-activation is possibly more energy efficient, as less muscle activity is needed. Our results suggest that training might improve the efficiency of balance control in children with cerebral palsy.

Similar underlying sensorimotor feedback pathways between rotational and translational perturbations might have facilitated the transfer between trained (i.e., rotational) and untrained (i.e., translational) perturbations [25]. Reciprocal inhibition, a mechanism that facilitates activation of the agonist muscle while suppressing antagonistic activity, is often reduced in children with cerebral palsy [30], [31], [32] and likely contributes to the excessive agonist-antagonist co-activation. It would therefore be interesting to investigate whether reciprocal inhibition is increased after perturbation training because a more mechanistic understanding of training-induced alterations in motor control can contribute to a better understanding of how training effects generalize to other functional movements, and can ultimately support personalized treatment selection that targets specific impairments in children with cerebral palsy.

High variability in the response to perturbation training was observed across our participants. Part of this variability might be due to the heterogeneity in the type and severity of their impairments including differences in motor subtype, severity, spasticity, and associated impairments. We included both unilaterally (N=6)- and bilaterally (N=8) affected children, children with and without plantarflexor spasticity (as measured by the MAS), and different secondary impairments. We were not able to attribute differences in co-contraction index to different MAS scores or to the involvement being unilateral vs. bilateral (**Appendix S5, figures S11-14**). This may indicate that commonly used clinical descriptors do not adequately capture the responsiveness to perturbation training. However, the limited sample size may also have reduced our ability to detect such associations.

We do not know whether the positive effects of perturbation training might persist after training and transfer to daily life function. Woollacott & Shumway Cook found improved muscle coordination one month after the end of the training intervention [8]. However, we did not include a retention session in our protocol. In addition, they found improved Gross Motor Function Measure (GMFM) for dimension D, which measures steady state balance control in children with CP, immediately after the end of the training intervention [7]. The GMFM is a clinical scale that is often used to score general gross motor function in children with cerebral palsy [33]. However, currently there are no tests within the GMFM that measure reactive balance control. As the ultimate purpose of balance training is to reduce the number of daily falls, future work might need to consider to test the retention and the effect on daily falls.

Some of the observed improvements in balance performance or muscle coordination may be attributed to test familiarization, as the participants underwent repeated translational perturbations during both pre- and post-measurements sessions. As a result of the training, children became familiar with the testing protocol, the testing environment, and the effect of platform movement, potentially reducing fear. However, test familiarization alone is unlikely to fully explain our findings, as improvements were higher in children with cerebral palsy than in typically developing children. This is in line with our expectations, given that the balance performance in the typically developing group was already high at baseline, leaving limited room for improvement. In addition, note that the trained rotational perturbations and the untrained translational perturbations were not entirely independent. The rotational perturbations used during training consisted of combined rotational and translational platform movement (**Figure 2**), meaning that participants were also exposed to a small translational component during training. Consequently, part of the observed improvement in the translational condition may reflect carry-over effects from this shared mechanical component.

Our limited sample might be biased towards children with limited impairments in reactive balance control. All recruited children were highly functional, as they were classified as GMFCS I or II and were able to walk at least 500m independently. In addition, most of the children with cerebral palsy were already able to suppress plantarflexor activity in time bin 3 from the rotational perturbations before training, while in a recent study on rotational perturbations in 20 children with cerebral palsy, almost all children were not able to suppress this unfavorable lateral gastrocnemius activity [13]. Furthermore, half of the children were already able to perform the highest levels of perturbations before training. Therefore, the high functionality of our measured children might limit the possible improvements. Further work should recruit more participants with different levels of involvement, as also existing literature only trained a very limited sample of children (N=2 and N=6) [8], [34].

Measuring muscle activity on two time points poses challenges when comparing the amplitude of muscle responses. For both sessions, we scaled the muscle activity to average EMG amplitude observed during overground walking in that particular session, as we do not expect substantial changes in average muscle activity during walking after training (gait speed was the same between sessions). However, we cannot exclude that scaling affected our results. If the training would have decreased muscle co-activation during walking as well, the average muscle activity during walking might be lower after training. This would lead to a lower scaling factor post-training compared to pre-training. Such lower scaling factor post-training would lead to higher scaled muscle activity and therefore the effect of training might have been underestimated. However, visual inspection of the walking trials did not reveal any changes in patterns of muscle activity. Therefore, we assume that the effect of scaling on our results is minimal. In addition, two EMG systems were used throughout the study because data was collected over a long time period (2021-2025) and during this period, the original system was replaced by a newer model. We do not expect this to have affected our findings as pre-and post-measurement sessions of each participant were always collected using the same EMG system.

Several important questions remain to be addressed in future studies. First, identification of the neural mechanisms underlying training-induced changes in muscle coordination could help to predict training responsiveness and support personalized treatment plans. Second, optimization of training intensity and volume and investigation of the long-term retention and functional relevance for daily life activities is needed. Finally, a larger cohort covering a broader spectrum of cerebral palsy severity is needed.

To conclude, we demonstrated that increased co-activation in children with cerebral palsy can be reduced through perturbation training. After training, children with cerebral palsy used balance control strategies that were more similar to those of typically developing children in both trained and untrained conditions, although muscle co-activation remained higher. It is promising that training effects were also observed in children older than 14, when balance control is fully developed in typically developing peers [35], [36]. It remains to be investigated whether the reduction in muscle co-activation after training generalizes beyond standing and reduces fall risk. The current lab-based setup is not suitable for clinical implementation but rotational perturbations can be administered with simpler devices making this a feasible intervention that is worthwhile to implement if the benefits for daily life functioning can be demonstrated.

## Data Availability

All data produced in the present study are available upon reasonable request to the authors.

## Data availability

The data supporting the conclusions of this article will be made available by the authors upon request, without undue reservation.

## Author contributions

J.W. and F.D.G conceived and designed research; J.W. performed experiments; J.W. analyzed data; J.W., A.V.C., K.D., and F.D.G interpreted results of experiments; J.W. and F.D.G. prepared figures; J.W. and F.D.G. drafted manuscript; J.W., A.V.C, K.D., and F.D.G edited and revised the manuscript; J.W., A.V.C., K.D., and F.D.G approved the final version of the manuscript.

## Acknowledgements

We thank all our participants for participating in this study.

