## Supplement data for "Rotational perturbation training improves muscle coordination during reactive standing balance in trained and untrained conditions in children with spastic cerebral palsy"

**SUPPLEMENTARY MATERIAL**

**S1. Training protocol**

Table S1: Overview of levels used during training sessions for all participants.

| | **Training 1** | **Training 2** | **Training 3** | **Training 4** | **Training 5** | **Training 6** | **Training 7** | **Training 8** | **Training 9** |
|---|---|---|---|---|---|---|---|---|---|
| **CP1** | L4-L5 | L4-L5 | L5-L6 | L5-L6 | L5-L6 | L5-L6 | L5-L6 | L5-L6 | L5-L6 |
| **CP2** | L3-L4 | L2-L3 | L2-L4 | L2-L3 | L3-L4 | L2-L5 | L2-L5 | L2-L6 | L3-L6 |
| **CP3** | L1-L2 | L1-L2 | L3-L3 | L3-L4 | L4-L5 | L5-L6 | L5-L6 | L5-L6 | L5-L6 |
| **CP4** | L1-L3 | L2-L4 | L3-L5 | L4-L6 | L5-L6 | L5-L6 | L5-L6 | L5-L6 | L5-L6 |
| **CP5** | L1-L4 | L2-L5 | L3-L5 | L3-L6 | L4-L6 | L5-L6 | L5-L6 | L2-L5 | L5-L6 |
| **CP6** | L1-L4 | L2-L4 | L3-L5 | L4-L5 | L4-L6 | L5-L6 | L2-L6 | L5-L6 | L5-L6 |
| **CP7** | L1-L1 | L1-L2 | L2-L3 | L3-L4 | L4-L5 | L5-L6 | L4-L6 | L5-L6 | L4-L6 |
| **CP8** | L1-L3 | L2-L4 | L3-L5 | L4-L6 | L5-L6 | L2-L6 | L3-L5 | L1-L4 | L5-L6 |
| **CP9** | L1-L3 | L2-L4 | L3-L5 | L4-L6 | L5-L6 | L2-L6 | L2-L5 | L1-L4 | L5-L6 |
| **CP10** | L1-L3 | L2-L4 | L3-L4 | L3-L5 | L4-L6 | L5-L6 | L2-L6 | L3-L5 | L5-L6 |
| **CP11** | L1-L3 | L2-L4 | L3-L5 | L4-L6 | L5-L6 | L2-L6 | L3-L5 | L1-L4 | L5-L6 |
| **CP12** | L1-L4 | L2-L5 | L3-L6 | L5-L6 | L2-L6 | L3-L5 | L1-L4 | L3-L6 | L5-L6 |
| **CP13** | L1-L4 | L2-L5 | L3-L6 | L5-L6 | L3-L6 | L3-L5 | L1-L4 | N/A | L5-L6 |
| **CP14** | L1-L4 | L2-L5 | L3-L6 | L5-L6 | L3-L6 | L2-L5 | L1-L4 | L3-L6 | L5-L6 |
| **TD1** | L5-L6 | L5-L6 | L5-L6 | L5-L6 | L5-L6 | L5-L6 | L5-L6 | L5-L6 | L5-L6 |
| **TD2** | L4-L5 | L5-L6 | L5-L6 | L5-L6 | L5-L6 | L5-L6 | L5-L6 | L5-L6 | L5-L6 |
| **TD3** | L5-L6 | L5-L6 | L5-L6 | L5-L6 | L5-L6 | L5-L6 | L5-L6 | L5-L6 | L5-L6 |
| **TD4** | L5-L6 | L5-L6 | L5-L6 | L5-L6 | L5-L6 | L5-L6 | L5-L6 | L5-L6 | L5-L6 |
| **TD5** | L1-L4 | L2-L5 | L3-L6 | L5-L6 | L1-L5 | L4-L6 | L5-L6 | L2-L6 | L5-L6 |
| **TD6** | L1-L4 | L2-L5 | L3-L6 | L5-L6 | L1-L5 | L4-L6 | L5-L6 | L2-L6 | L5-L6 |
| **TD7** | L1-L4 | L2-L5 | L3-L6 | L5-L6 | L1-L5 | L4-L6 | L5-L6 | L2-L6 | L5-L6 |
| **TD8** | L1-L4 | L2-L5 | L3-L6 | L5-L6 | L2-L6 | L3-L5 | L1-L4 | L4-L6 | L5-L6 |
| **TD9** | L1-L4 | L2-L5 | L3-L6 | L5-L6 | N/A | L3-L5 | L1-L4 | L3-L6 | L5-L6 |
| **TD10** | L1-L4 | L2-L5 | L3-L6 | L5-L6 | L2-L6 | L3-L5 | L1-L4 | L4-L6 | L5-L6 |

**S2. Marker placement**

Table S2: Overview of marker placement during pre- and post-measurement sessions.

| Segment | Marker | | Position |
|---|---|---|---|
| Torso | Acromion | (L + R) | Most superior point of the scapula |
| | Sternum | | Upper edge of sternum |
| Pelvis | Spina Iliaca Anterior Superior | (L + R) | Most pronounced part of SIAS |
| | Spina iliace Posterior Superior | (L + R) | Most pronounced part of SIPS |
| | Crista Iliaca | (L + R) | Superior border of ilium |
| Thigh | Greater trochanter | (L + R) | Most lateral part of the hip |
| | Cluster thigh | (L + R) | 3 markers in triangle on thigh |
| | Lateral epicondyle of the femur | (L + R) | Most pronounced part of the epicondyle |
| | Medial epicondyle of the femur | (L + R) | Most pronounced part of the epicondyle |
| Shank | Cluster shank | (L + R) | 3 markers in triangle on shank |
| | Lateral malleolus | (L + R) | Most pronounced part of mallelolus |
| | Medial malleolus | (L + R) | Most pronounced part of malleolus |
| Foot | Heel | (L + R) | Dorsal part of the calcaneus |
| | Lateral calcaneus | (L + R) | Lateral aspect of calcaneus, equidistant from heel |
| | Medial calcaneus | (L + R) | Medial aspect of calcaneus, equidistant from heel |
| | 1st metatarsal head | (L + R) | On top of the 1st metatarsal head |
| | 5th metatarsal head | (L + R) | On top of the 5th metatarsal head |
| Arm | Lateral epicondyle of the humerus | (L + R) | Most pronounced part of the epicondyle |
| | Medial epicondyle of the femur | (L + R) | Most pronounced part of the epicondyle |
| | Ulna | (L + R) | Styloid processus ulna |
| | Radius | (L + R) | Distal end of radius |
| | Finger | (L + R) | Head of 3th metacarpal bone |

**S3. Co-contraction index pre vs. post**

Table S3: Statistical output of co-contraction index pre vs. post training for children with CP and TD children

| | | Cerebral palsy | | Typically developing | |
|---|---|---|---|---|---|
| | | *F- value* | *P* | *F- value* | *P* |
| **TO** | LG-TA | 2.25 | **0.046** | 1.75 | 0.119 |
| | MG-TA | 1.40 | 0.224 | 0.30 | 0.934 |
| | SOL-TA | 2.49 | **0.027** | 3.38 | **0.005** |
| **BW** | LG-TA | 2.52 | **0.027** | 0.21 | 0.972 |
| | MG-TA | 3.14 | **0.008** | 0.07 | 0.999 |
| | SOL-TA | 2.71 | **0.018** | 0.29 | 0.939 |

TO = toe-up rotational perturbation; BW= backward translational perturbation; LG = lateral gastrocnemius; MG = medial gastrocnemius; SOL = soleus; TA = tibialis anterior.

Table S4: Statistical output of co-contraction index CP vs. TD

| | | CP vs. TD | |
|---|---|---|---|
| | | *F-value* | *P* |
| **TO** | LG-TA | 3.12 | **0.006** |
| | MG-TA | 6.48 | **<0.0001** |
| | SOL-TA | 8.58 | **< 0.0001** |
| **BW** | LG-TA | 3.92 | **0.001** |
| | MG-TA | 4.04 | **0.001** |
| | SOL-TA | 7.08 | **<0.001** |

TO = toe-up rotational perturbation; BW= backward translational perturbation; LG = lateral gastrocnemius; MG = medial gastrocnemius; SOL = soleus; TA = tibialis anterior.

S3.1 Rotational perturbations

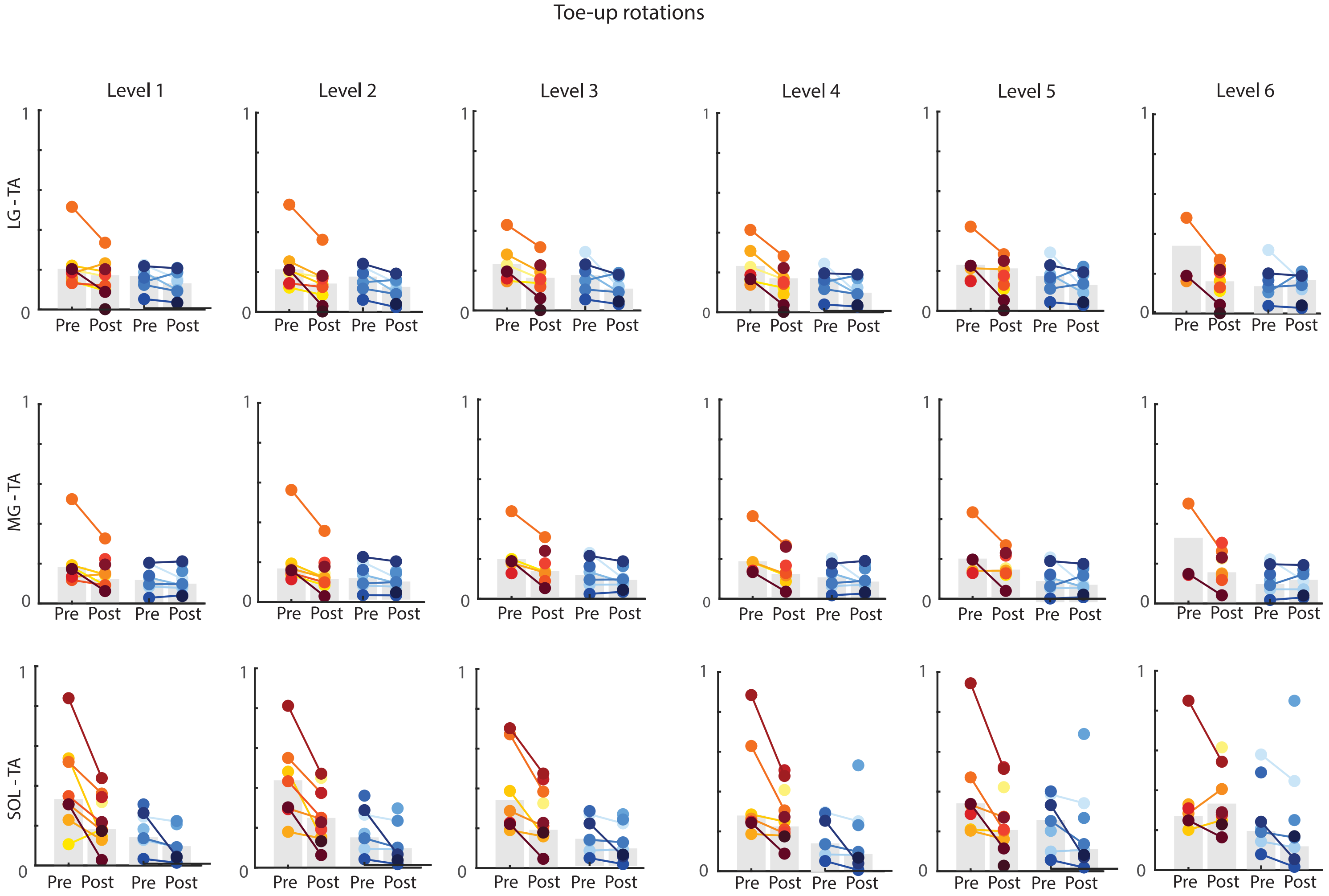

Figure S1: Co-contraction index between plantarflexors (lateral gastrocnemius (LG), medial gastrocnemius (MG), soleus (SOL)) and tibialis anterior (TA) for the toe-up rotational perturbations. Children with cerebral palsy (CP) in red/yellow colors (left side of each subplot) and typically developing (TD) children in blue (right side of each subplot).

3.2 Translational perturbations

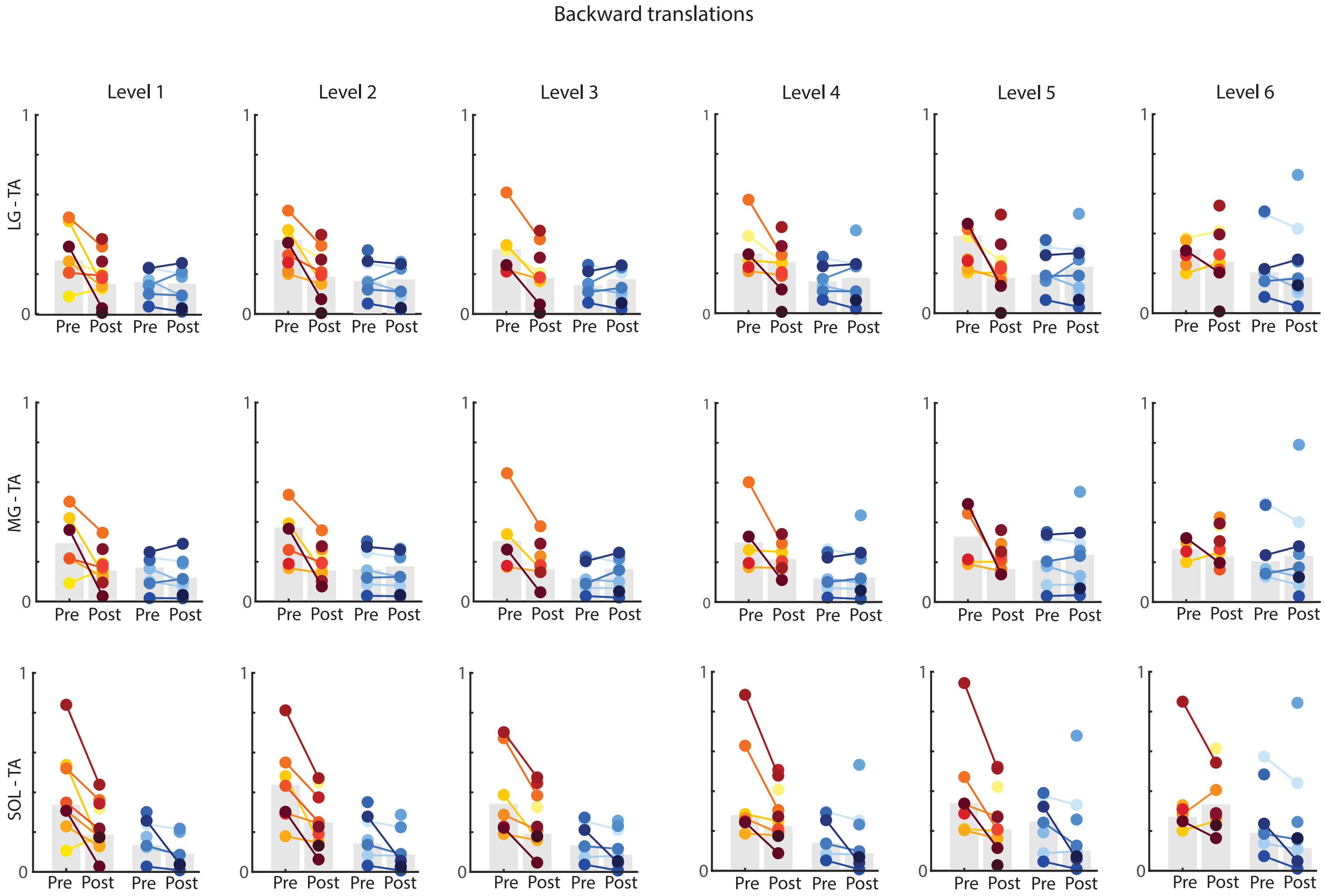

Figure S2: Co-contraction index between plantarflexors (lateral gastrocnemius (LG), medial gastrocnemius (MG), soleus (SOL)) and tibialis anterior (TA) for the backward translational perturbations. Children with cerebral palsy (CP) in red/yellow colors (left side of each subplot) and typically developing (TD) children in blue (right side of each subplot).

**S4. EMG Zones pre vs. Post**

Table S5: Statistical output for average muscle activity (zones) pre vs. post training for children with CP and TD children

| | | **Cerebral palsy** | | | | | | **Typically developing** | | | | | |
|---|---|---|---|---|---|---|---|---|---|---|---|---|---|
| | | **Zone 1** | | **Zone 2** | | **Zone 3** | | **Zone 1** | | **Zone 2** | | **Zone 3** | |
| | | *F- value* | *P* | *F- value* | *P* | *F- value* | *P* | *F- value* | *P* | *F- value* | *P* | *F- value* | *P* |
| **TO** | LG | 0.990 | 0.436 | 3.640 | **0.003** | 6.860 | **<0.0001** | 0.790 | 0.579 | 2.990 | **0.011** | 1.780 | 0.112 |
| | MG | 0.780 | 0.589 | 1.950 | 0.081 | 2.670 | **0.019** | 0.670 | 0.675 | 7.920 | **<0.0001** | 2.450 | **0.031** |
| | SOL | 0.910 | 0.489 | 3.650 | **0.002** | 2.160 | **0.050** | 0.920 | 0.483 | 1.340 | 0.250 | 1.270 | 0.278 |
| | TA | 1.690 | 0.130 | 0.850 | 0.533 | 1.500 | 0.185 | 0.550 | 0.766 | 3.030 | **0.009** | 5.690 | **<0.0001** |
| **BW** | LG | 0.470 | 0.830 | 0.540 | 0.780 | 1.050 | 0.394 | 1.890 | 0.091 | 0.380 | 0.891 | 0.550 | 0.770 |
| | MG | 1.060 | 0.393 | 0.620 | 0.716 | 4.500 | **0.001** | 1.230 | 0.297 | 1.360 | 0.241 | 1.960 | 0.080 |
| | SOL | 0.680 | 0.664 | 1.300 | 0.264 | 2.990 | **0.010** | 0.310 | 0.929 | 0.470 | 0.827 | 1.130 | 0.353 |
| | TA | 0.740 | 0.616 | 1.980 | 0.074 | 5.080 | **0.000** | 0.810 | 0.566 | 1.260 | 0.284 | 1.640 | 0.145 |

TO = toe-up rotational perturbations; BW = backward translational perturbations; LG = lateral gastrocnemius; MG = medial gastrocnemius; SOL = soleus; TA = tibialis anterior.

### S4.1 Rotational perturbations

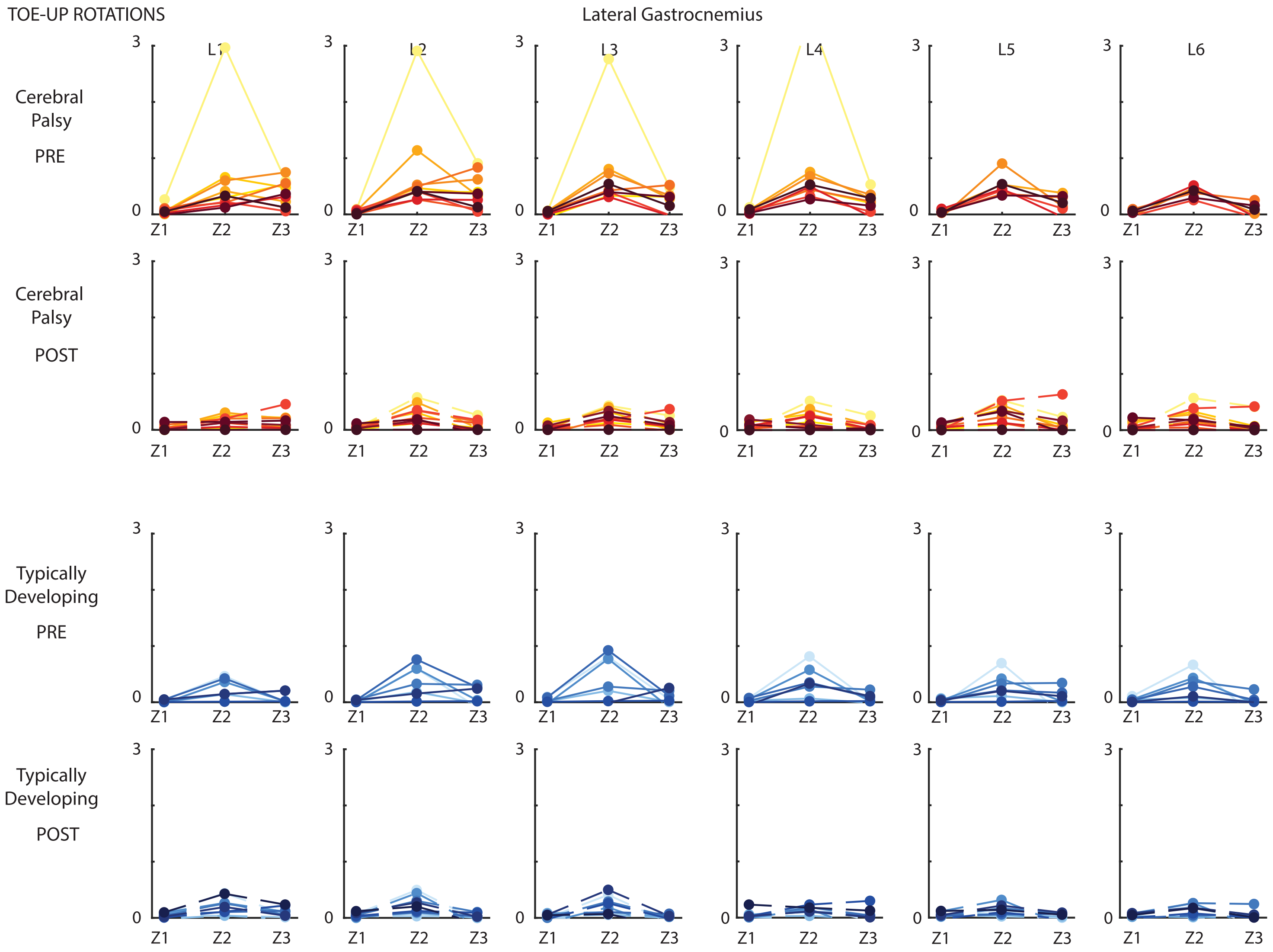

Figure S3: Average normalized EMG for the lateral gastrocnemius for three time bins (Z1-Z3) for all levels (L1-L6) the rotational perturbations pre (full line)- and post (dotted line) intervention. Children with cerebral palsy (CP) in red/yellow colors (top two panels) and typically developing (TD) children in blue (bottom two panels).

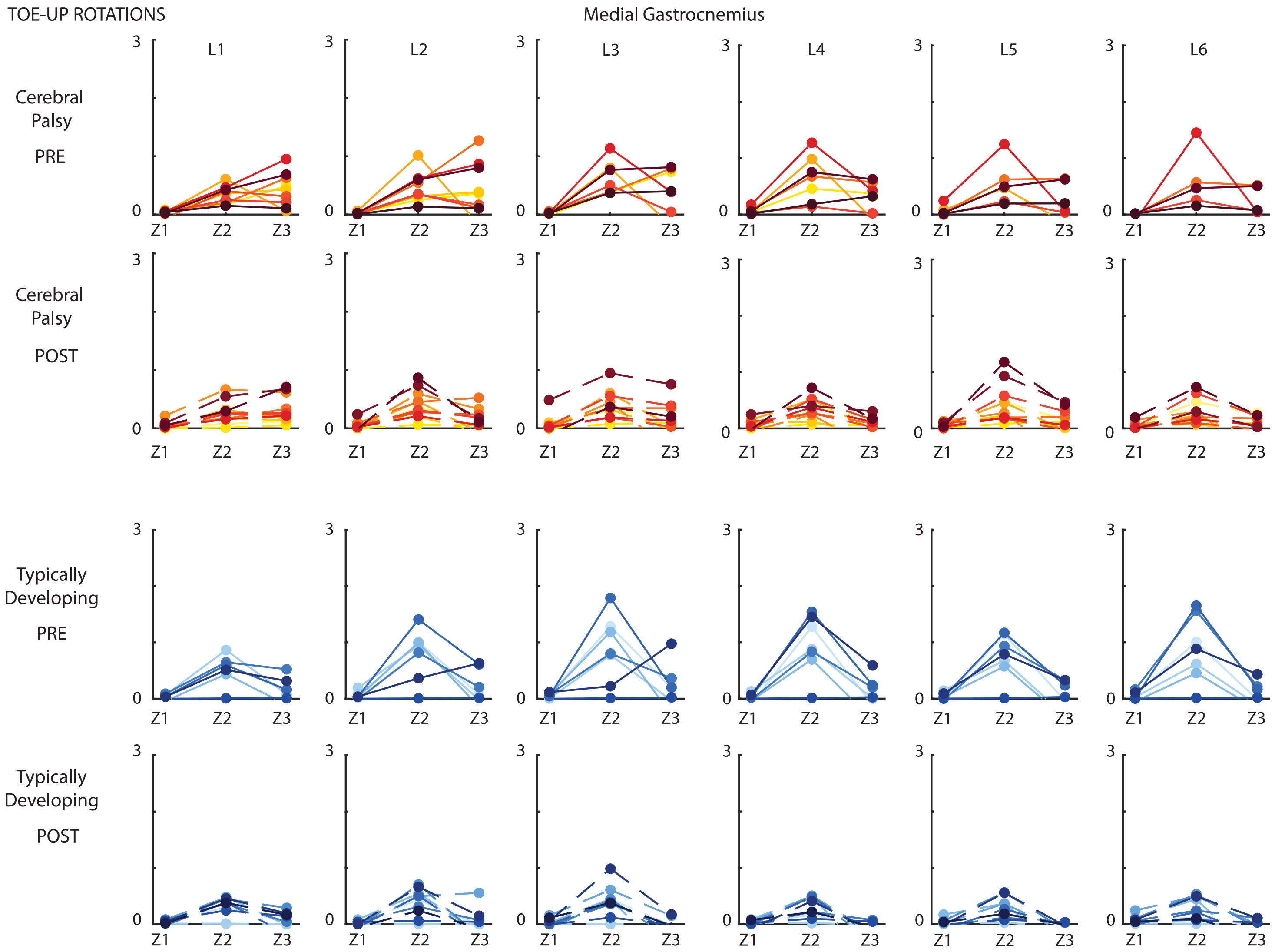

Figure S4: Average normalized EMG for the medial gastrocnemius for three time bins (Z1-Z3) for all levels (L1-L6) the rotational perturbations pre (full line)- and post (dotted line) intervention. Children with cerebral palsy (CP) in red/yellow colors (top two panels) and typically developing (TD) children in blue (bottom two panels).

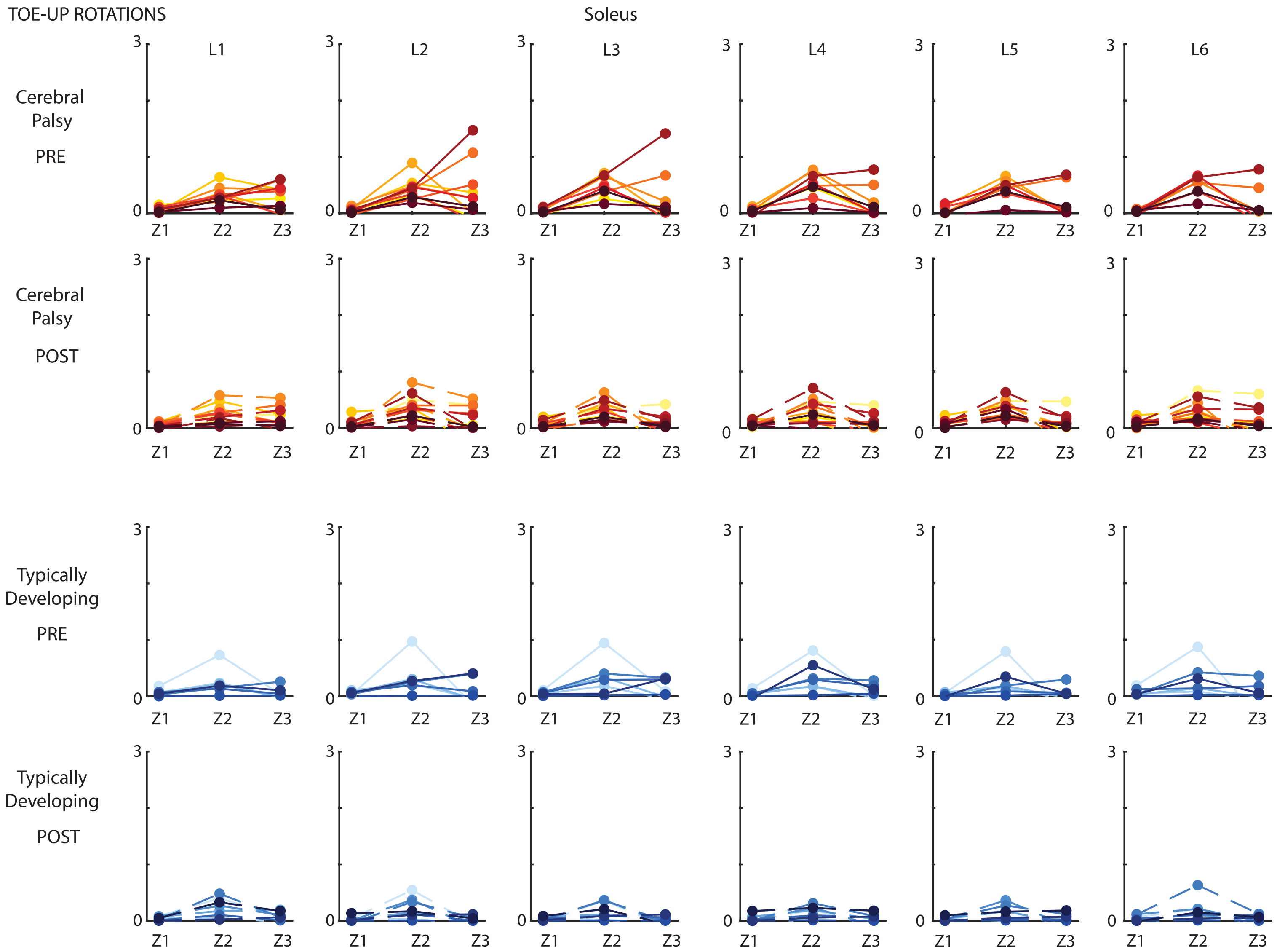

Figure S5: Average normalized EMG for the soleus for three time bins (Z1-Z3) for all levels (L1-L6) the rotational perturbations pre (full line)- and post (dotted line) intervention. Children with cerebral palsy (CP) in red/yellow colors (top two panels) and typically developing (TD) children in blue (bottom two panels).

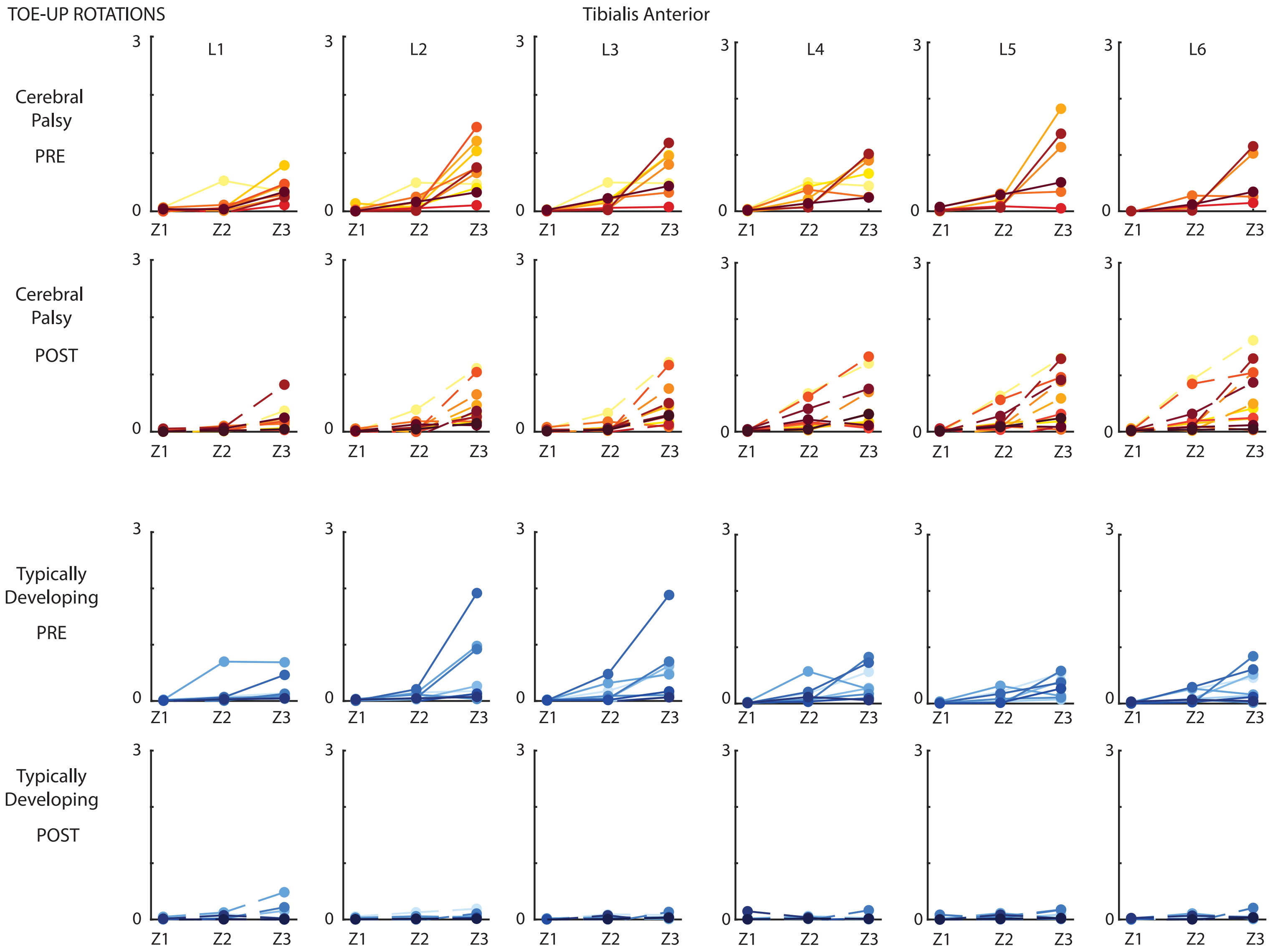

Figure S6: Average normalized EMG for the soleus for three time bins (Z1-Z3) for all levels (L1-L6) the rotational perturbations pre (full line)- and post (dotted line) intervention. Children with cerebral palsy (CP) in red/yellow colors (top two panels) and typically developing (TD) children in blue (bottom two panels).

S4.2 Backward translations

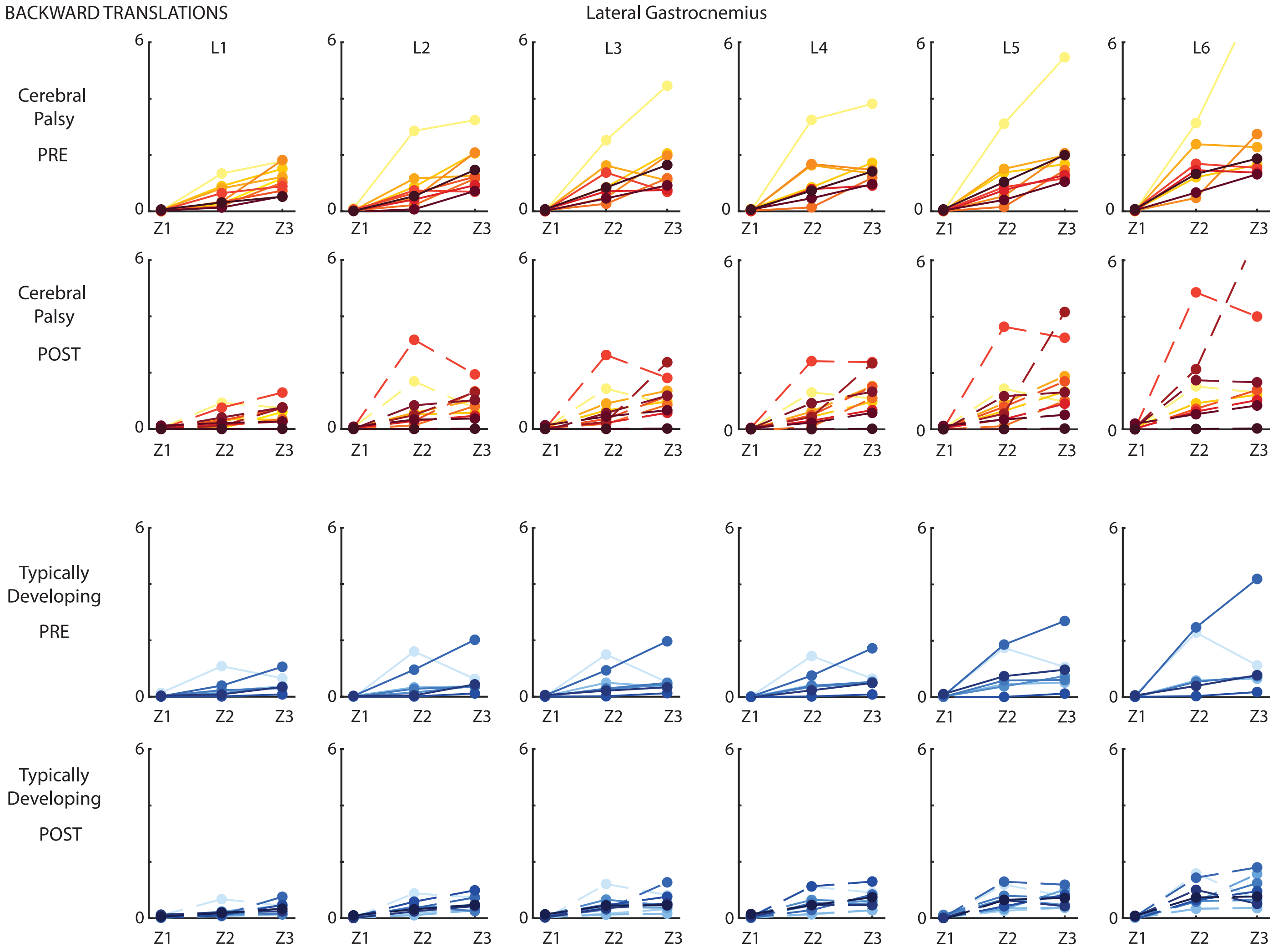

Figure S7: Average normalized EMG for the lateral gastrocnemius for three time bins (Z1-Z3) for all levels (L1-L6) the translational perturbations pre (full line)- and post (dotted line) intervention. Children with cerebral palsy (CP) in red/yellow colors (top two panels) and typically developing (TD) children in blue (bottom two panels).

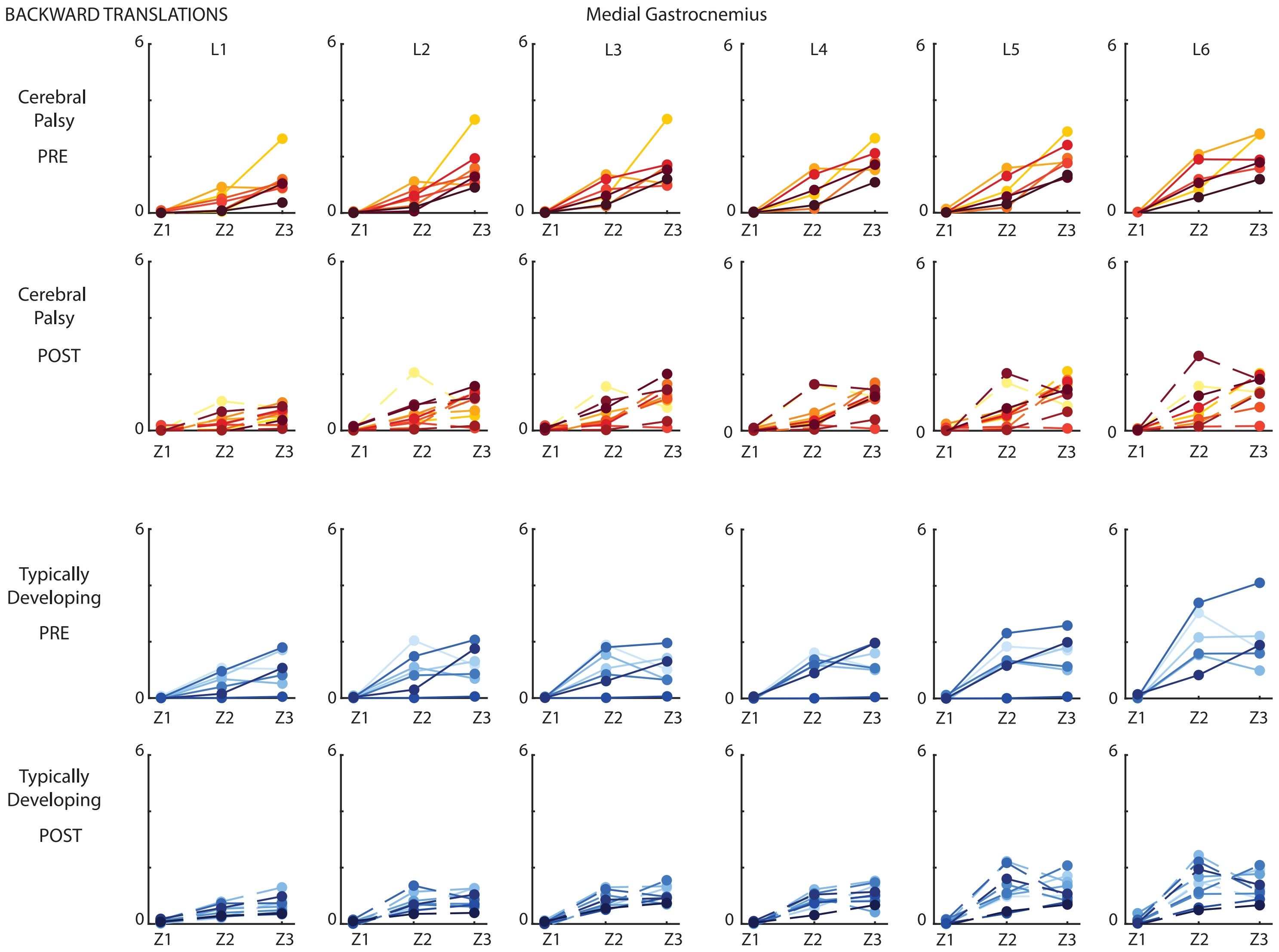

Figure S8: Average normalized EMG for the medial gastrocnemius for three time bins (Z1-Z3) for all levels (L1-L6) the translational perturbations pre (full line)- and post (dotted line) intervention. Children with cerebral palsy (CP) in red/yellow colors (top two panels) and typically developing (TD) children in blue (bottom two panels).

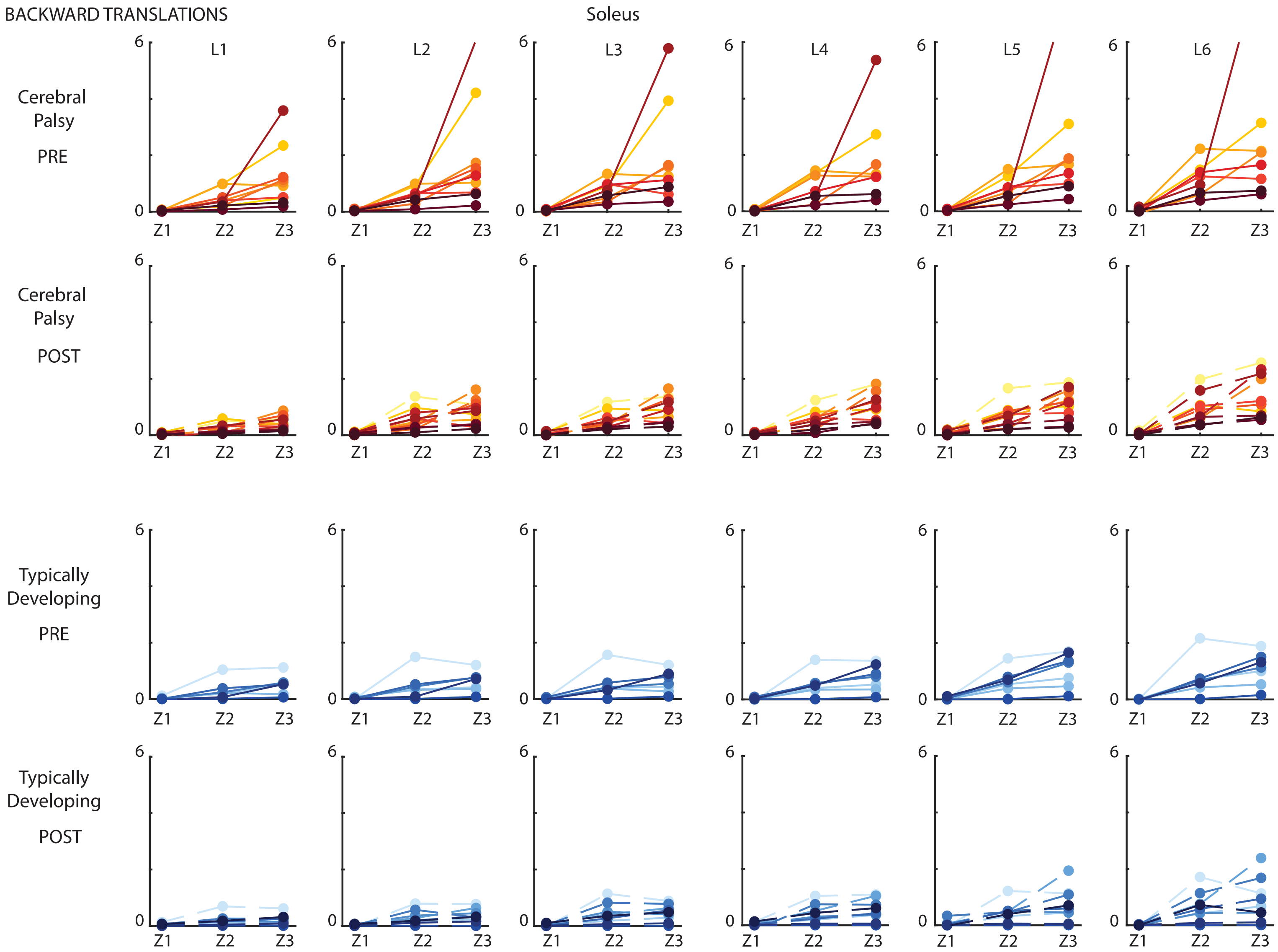

Figure S9: Average normalized EMG for the soleus for three time bins (Z1-Z3) for all levels (L1-L6) the translational perturbations pre (full line)- and post (dotted line) intervention. Children with cerebral palsy (CP) in red/yellow colors (top two panels) and typically developing (TD) children in blue (bottom two panels).

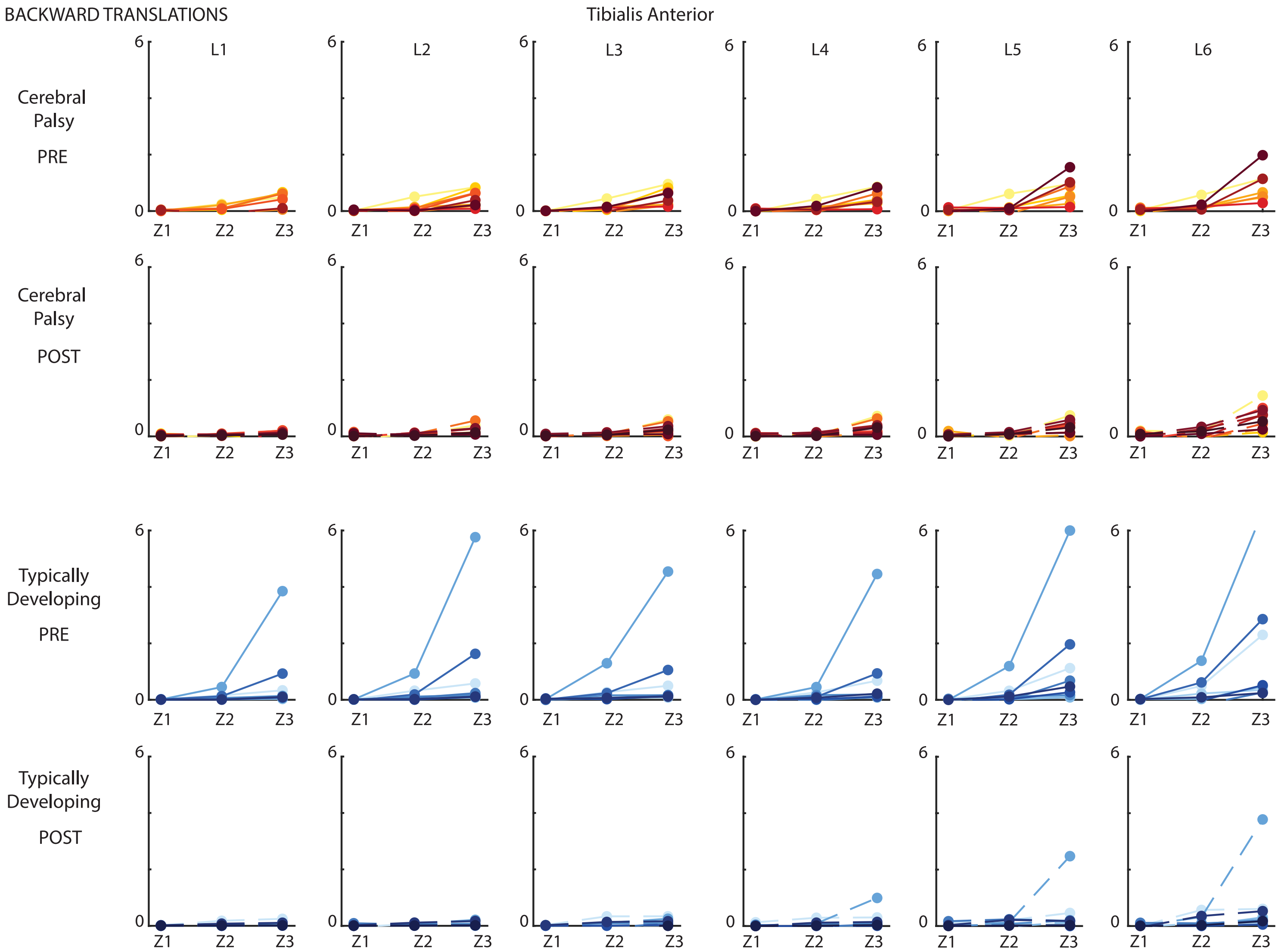

Figure S10: Average normalized EMG for the tibialis anterior for three time bins (Z1-Z3) for all levels (L1-L6) the translational perturbations pre (full line)- and post (dotted line) intervention. Children with cerebral palsy (CP) in red/yellow colors (top two panels) and typically developing (TD) children in blue (bottom two panels).

**S5. Demographic influence on CCI**

S5.1 Modified Ashworth Scale

### S5.1.1 Rotational Perturbations

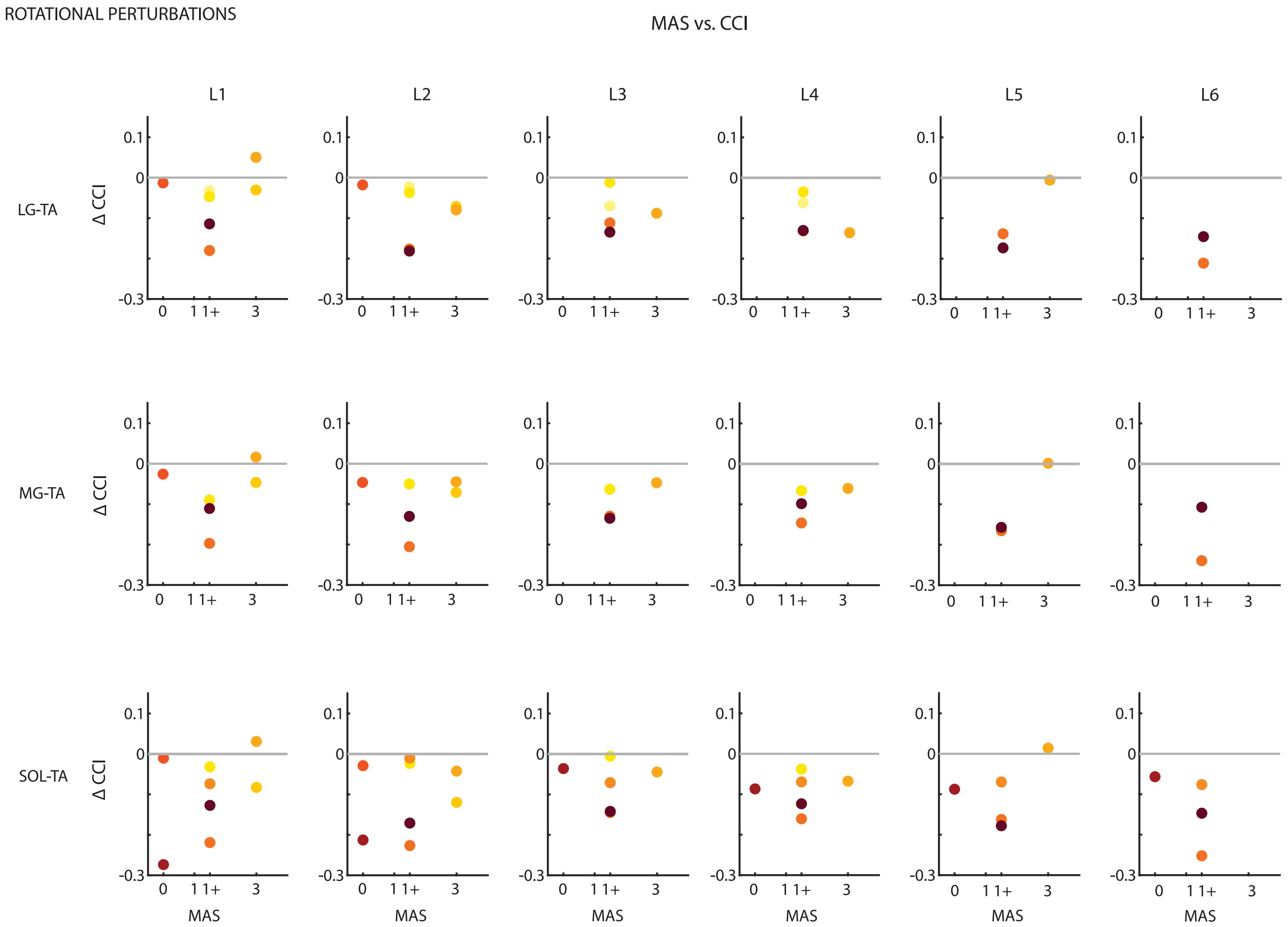

Figure S11: Delta (post-pre) co-contraction index (CCI) for all levels (L1-L6) of rotational perturbations to explore the difference between participants with different Modified Ashworth Scores (MAS). Each color represents one child with cerebral palsy.

### 5.1.2 Translational Perturbations

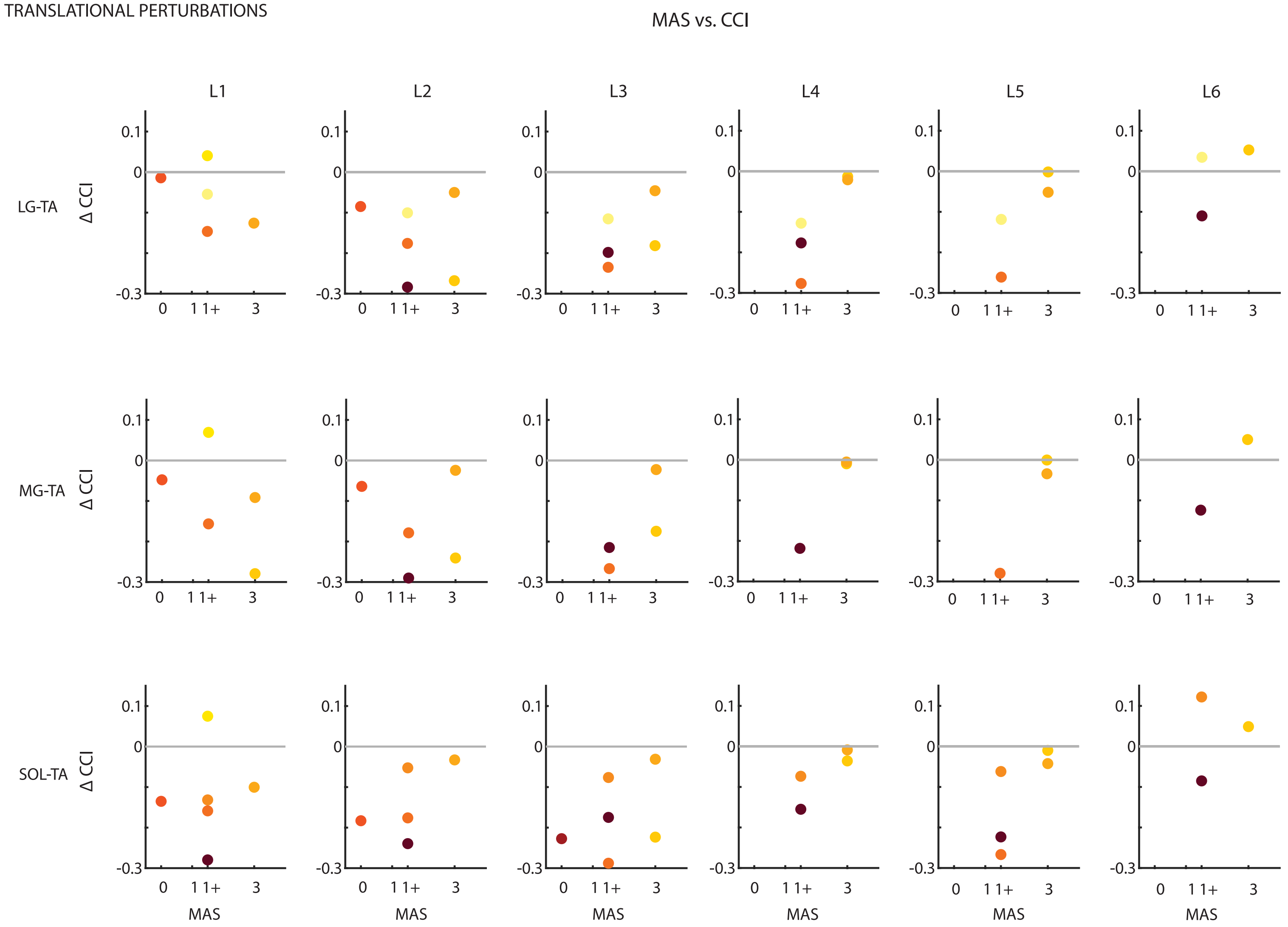

Figure S12: Delta (pre-post) co-contraction index (CCI) for all levels (L1-L6) of translational perturbations to explore the difference between participants with different Modified Ashworth Scores (MAS). Each color represents one child with cerebral palsy.

S5.2 Hemiplegic vs. Diplegic

S5.2.1 Rotational perturbations

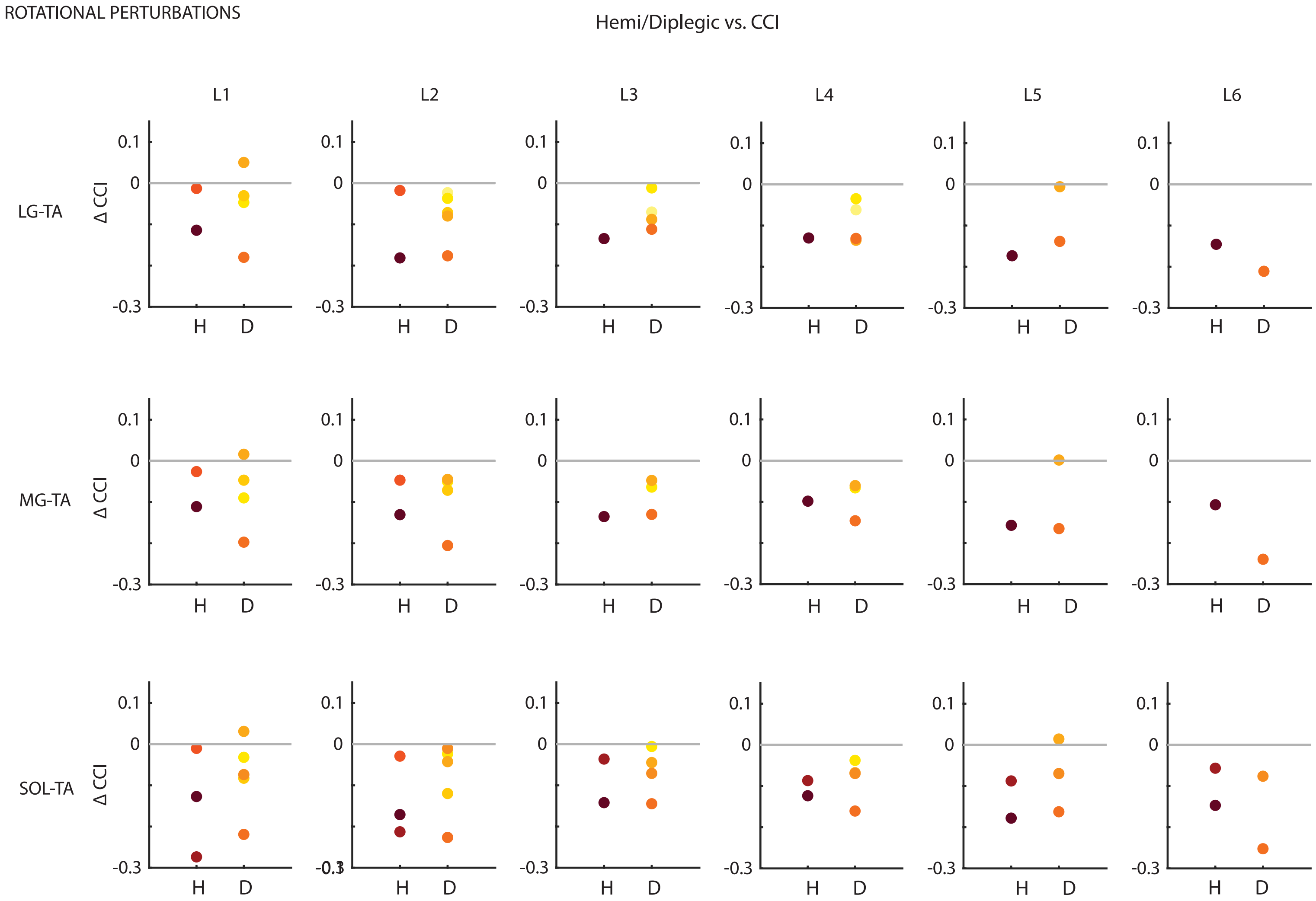

Figure S13: Delta (pre-post) co-contraction index (CCI) for all levels (L1-L6) of rotational perturbations to explore the difference between participants with Hemiplegic (H) vs. Diplegic (D) involvement. Each color represents one child with cerebral palsy.

#### S5.2.2 Translational perturbations

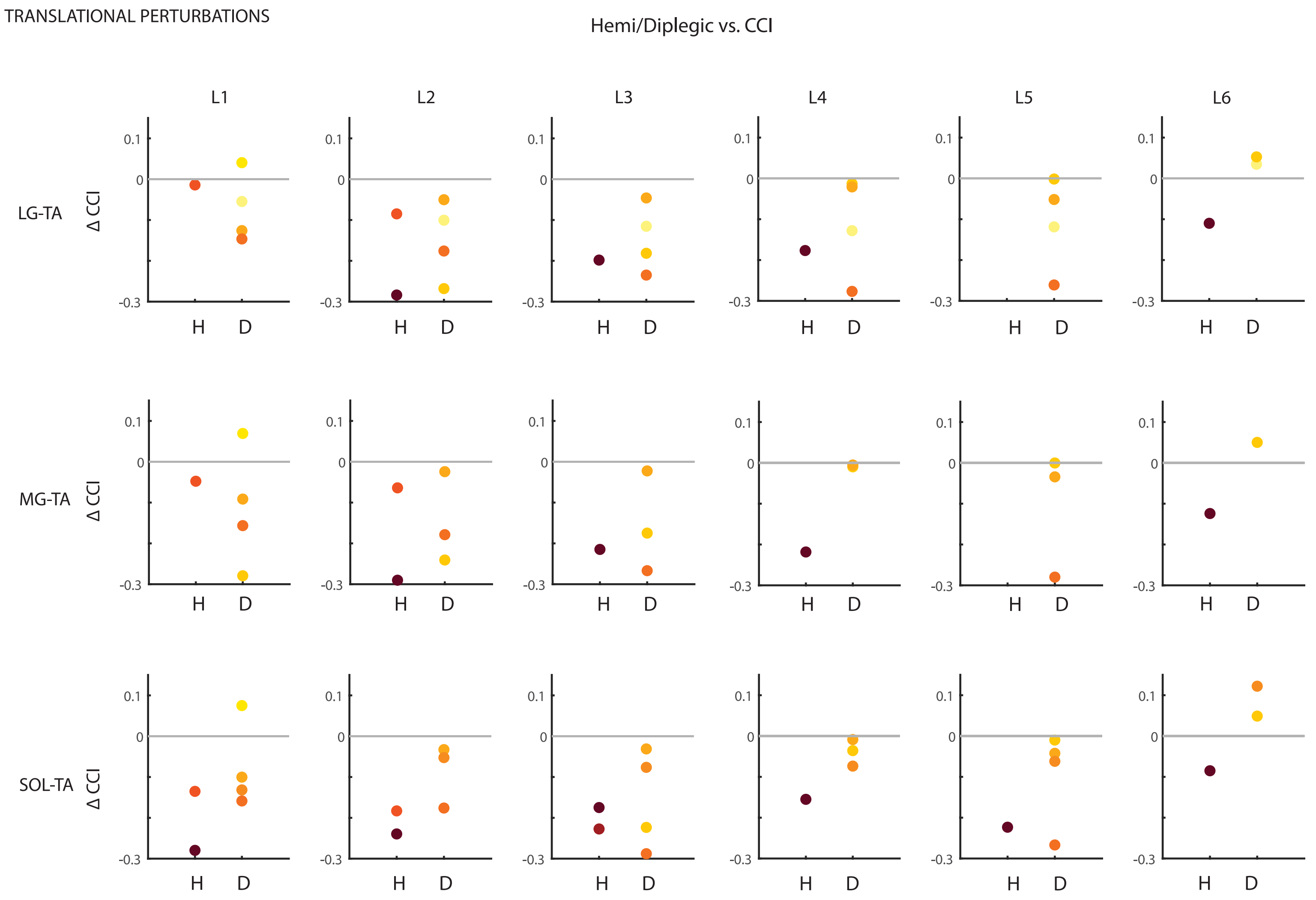

Figure S14: Delta (pre-post) co-contraction index (CCI) for all levels (L1-L6) of translational perturbations to explore the difference between participants with Hemiplegic (H) vs. Diplegic (D) involvement. Each color represents one child with cerebral palsy.
